# SARS-CoV-2 infects, replicates, elevates angiotensin II and activates immune cells in human testes

**DOI:** 10.1101/2022.02.05.22270327

**Authors:** Guilherme M.J. Costa, Samyra M.S.N. Lacerda, André F.A. Figueiredo, Natália T. Wnuk, Marcos R. G. Brener, Gabriel H. Campolina-Silva, Andrea Kauffmann-Zeh, Lucila GG Pacifico, Alice F. Versiani, Lídia M. Andrade, Maísa M. Antunes, Fernanda R. Souza, Geovanni D. Cassali, André L. Caldeira-Brant, Hélio Chiarini-Garcia, Vivian V. Costa, Flavio G. da Fonseca, Maurício L. Nogueira, Guilherme R. F. Campos, Lucas M. Kangussu, Estefânia M. N. Martins, Loudiana M. Antonio, Cintia Bittar, Paula Rahal, Renato S. Aguiar, Bárbara P. Mendes, Marcela S. Procópio, Thiago P. Furtado, Yuri L Guimaraes, Gustavo B Menezes, Ana Martinez-Marchal, Miguel Brieno-Enriquez, Kyle E. Orwig, Marcelo H. Furtado

## Abstract

Although much has been published since the first cases of COVID-19, there remain unanswered questions regarding SARS-CoV-2 impact on testes and the potential consequences for reproductive health. We investigated testicular alterations in deceased COVID-19-patients, the precise location of the virus, its replicative activity, and the molecules involved in the pathogenesis. We found that SARS-CoV-2 testicular tropism is higher than previously thought and that reliable viral detection in the testis requires sensitive nanosensoring or RT-qPCR using a specific methodology. Macrophages and spermatogonial cells are the main SARS-CoV-2 lodging sites and where new virions form inside the Endoplasmic Reticulum Golgi Intermediate Complex. Moreover, we showed infiltrative infected monocytes migrating into the testicular parenchyma. SARS-CoV-2 maintains its replicative and infective abilities long after the patient’s infection, suggesting that the testes may serve as a viral sanctuary. Further, infected testes show thickening of the tunica propria, germ cell apoptosis, Sertoli cell barrier loss, evident hemorrhage, angiogenesis, Leydig cell inhibition, inflammation, and fibrosis. Finally, our findings indicate that high angiotensin II levels and activation of mast cells and macrophages may be critical for testicular pathogenesis. Importantly, our data suggest that patients who become critically ill exhibit severe damages and may harbor the active virus in testes.

## INITRODUCTION

Since the testis displays one of the highest expressions of Angiotensin Converting Enzyme 2 (ACE2) receptors, which mediate the cellular entry of SARS-CoV-2, and current data suggest that men are more affected than women(1), deep testicular evaluations of patients affected by COVID-19 is imperative. Previous studies present discordant results concerning SARS-CoV-2 detection in testicular parenchyma through RT-PCR (2-5) and the extent of testicular damage caused by the virus(5). Moreover, many questions about viral infection remain unexplored, such as the viral replication, route of infection, and the identity of infected cells.

Although some testicular alterations promoted by SARS-CoV-2 infection were previously demonstrated(2-5), the players of testicular pathogenesis in COVID-19 remain unknown. Thus, studying the cellular, enzymatic, hormonal, and critical gene alterations in the testes of COVID-19 patients should contribute to a better understanding of SARS-CoV-2 biology and its possible impact on testes and male fertility. Recently, Edenfield and Easley(6) published a perspective article in Nature Reviews Urology urging the need to unveil the potential mechanisms involving SARS-CoV-2’s entry and pathophysiological effects on affected subjects.

We used different methods to detect SARS-CoV-2 in the testis parenchyma of patients deceased with COVID-19 and investigated the virus infective and replicative capacities. We also revealed the cellular and molecular alterations in human testicular pathophysiology and correlated the findings with the patients’ clinical data, unveiling potential mechanisms underlying the observed alterations

## MATERIAL AND METHODS

### COVID-19 PATIENTS

In 2021, we enrolled 11 non-vaccinated male patients deceased from COVID-19 complications, confirmed by SARS-CoV-2 RT-qPCR performed during their hospital stay, initially admitted in hospitals of Belo Horizonte, Brazil. All 11 patients were admitted to the ICU due to severe pulmonary symptoms (Table 1). The standard treatment at ICU included antibiotics, antimycotics, sedatives, muscular relaxants, analgesics, antihypertensives, inotropes, vasopressors agents, invasive mechanical ventilation, and hemodialysis. The Research Ethics Committee of the Mater Dei Hospital and the National Research Ethics Committee (CONEP) approved this investigation under the number CAAE: 30999320.1.0000.5128.

**Table 1-.**
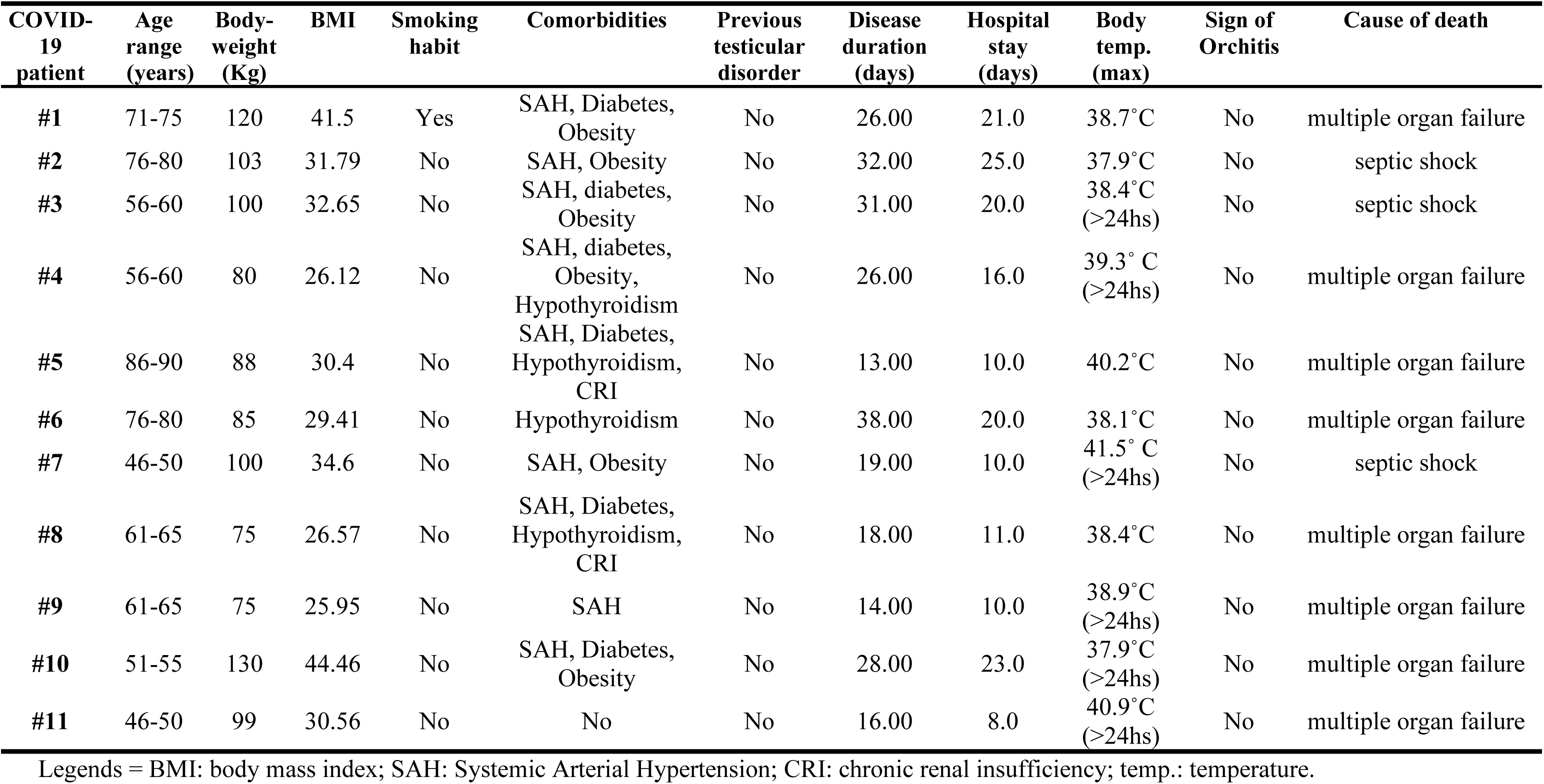
Clinical features of 11 COVID-19 patients.

| COVID-19 patient | Age range (years) | Body-weight (Kg) | BMI | Smoking habit | Comorbidities | Previous testicular disorder | Disease duration (days) | Hospital stay (days) | Body temp. (max) | Sign of Orchitis | Cause of death |
| --- | --- | --- | --- | --- | --- | --- | --- | --- | --- | --- | --- |
| #1 | 71-75 | 120 | 41.5 | Yes | SAH, Diabetes, Obesity | No | 26.00 | 21.0 | 38.7°C | No | multiple organ failure |
| #2 | 76-80 | 103 | 31.79 | No | SAH, Obesity | No | 32.00 | 25.0 | 37.9°C | No | septic shock |
| #3 | 56-60 | 100 | 32.65 | No | SAH, diabetes, Obesity | No | 31.00 | 20.0 | 38.4°C (>24hs) | No | septic shock |
| #4 | 56-60 | 80 | 26.12 | No | SAH, diabetes, Obesity, Hypothyroidism | No | 26.00 | 16.0 | 39.3° C (>24hs) | No | multiple organ failure |
| #5 | 86-90 | 88 | 30.4 | No | SAH, Diabetes, Hypothyroidism, CRI | No | 13.00 | 10.0 | 40.2°C | No | multiple organ failure |
| #6 | 76-80 | 85 | 29.41 | No | Hypothyroidism | No | 38.00 | 20.0 | 38.1°C | No | multiple organ failure |
| #7 | 46-50 | 100 | 34.6 | No | SAH, Obesity | No | 19.00 | 10.0 | 41.5° C (>24hs) | No | septic shock |
| #8 | 61-65 | 75 | 26.57 | No | SAH, Diabetes, Hypothyroidism, CRI | No | 18.00 | 11.0 | 38.4°C | No | multiple organ failure |
| #9 | 61-65 | 75 | 25.95 | No | SAH | No | 14.00 | 10.0 | 38.9°C (>24hs) | No | multiple organ failure |
| #10 | 51-55 | 130 | 44.46 | No | SAH, Diabetes, Obesity | No | 28.00 | 23.0 | 37.9°C (>24hs) | No | multiple organ failure |
| #11 | 46-50 | 99 | 30.56 | No | No | No | 16.00 | 8.0 | 40.9°C (>24hs) | No | multiple organ failure |

Legends = BMI: body mass index; SAH: Systemic Arterial Hypertension; CRI: chronic renal insufficiency; temp.: temperature.

Postmortem collection of both testicles was performed after a legally responsible family member signed an informed consent document. Testicles were collected through an incision on the median raphe of the scrotum. Two authors (MHF and YLG) collected all testicles no later than three hours after the patients’ death. To perform viral and testicular genetic studies, fragments of testicular parenchyma were sampled and immersed into RNAlater® solution (Sigma-Aldrich). To investigate the viral replicative activity and testosterone and angiotensin levels, testis fragments were also sampled and then snap-frozen in liquid nitrogen. The remaining testicular halves were immersed in different fixatives, such as Paraformaldehyde 4%, Bouin, Methacarn, and Glutaraldehyde 4%. Testis fragments were embedded in Methacrylate, Epon 812 resin, and Paraplast® for histological, transmission electron microscopy (TEM), and immunohistochemistry analyses.

### CONTROL PATIENTS

The Control group was composed of six patients who underwent orchiectomy due to prostate cancer suspicion. These patients did not go through any treatment at the time of orchiectomy. Moreover, they exhibited normal spermatogenesis in seminiferous tubules. The age and hormonal levels of these patients are presented in Supplemental Table 1. Testicular fragments were obtained after the study was approved by the Ethics Committee in Research of the Universidade Federal de Minas Gerais COEP/UFMG (COEP ETIC n°117/07). All patients signed the informed consent. Testicular samples were placed in liquid nitrogen, embedded in methacrylate, Epon, paraplast®, and conserved in RNAlatter. These samples were used for TEM, histological, hormonal, and molecular analyses. The mean age of patients was 58 years old, ranging from 46 to 65 years old.

### DETECTION OF SARS-CoV-2 IN TESTIS TISSUE GENETIC ASSAYS

#### SARS-CoV-2 detection using standard RT-qPCR

RNA of samples was extracted according to the protocol specified by the extraction kit. The following kit was used: QIAamp® Viral RNA Mini Kit. The samples were stored in an ultra-freezer at -80 °C for preservation. The collected samples were tested for the presence of SARS-CoV-2 viral RNA by RT-qPCR with primers to amplify the envelope (E) gene in addition to an endogenous control, the human transcript of the gene for RNAseP (RNP)(7). Samples were considered positives with a cycle threshold (CT) ≤ 40.

#### RT-qPCR using specific viral primers

Testes samples were macerated and submitted to RNA extraction using the Viral RNA Kit (Zymo Research, USA), following the manufacturer’s protocol. A two-step RT-qPCR approach was performed to optimize the detection of the viral RNA in the tissue samples and to avoid the possible influence of host cellular RNA. The obtained RNA was first submitted to cDNA synthesis using the CDC’s SARS-CoV-2 specific reverse primer 2019-nCoV_N1-R (TCT GGT TAC TGC CAG TTG AAT CTG) and the SuperScript™ III First-Strand Synthesis System (Invitrogen, Brazil).

The viral cDNAs were then amplified in a qPCR reaction using the GoTaq qPCR Master Mix (Promega, USA). We used both N1 primers from CDC’s SARS-CoV-2 detection protocol (2019-nCoV_N1-F: GAC CCC AAA ATC AGC GAA AT; 2019-nCoV_N1-R: TCT GGT TAC TGC CAG TTG AAT CTG) and followed the cycling recommendation indicated by the enzyme’s supplier in a QuantStudio 3 Real-Time PCR System (Applied Biosystems, USA). To normalize the results, the same process was performed to amplify the human β-actin control.

### PROTEIN ASSAYS

#### Nanosensor

Gold nanorods (GNRs) were synthesized by the seed-mediated growth method as previously described(8). The nanoparticles contained an average aspect ratio of 10 x 38 nm and a light absorbance peak of 713nm. They were covalently functionalized with the polyclonal antibody anti-Spike protein (Rhea Biotech, Brazil) and a polyclonal antibody anti-Nucleocapsid protein (CTVacinas, Brazil) through a carbodiimide-activated amidation reaction. The binding between the gold surface and the antibodies was mediated by adding a capping layer formed by α-lipoic acid. A 2mM α-lipoic acid solution (LA; Sigma Aldrich, USA) in ethanol was added to the GNR suspension (0.039mg/mL). These suspensions were exposed to an ultrasonic bath (UNIQUE model U5C1850, 154W, 25KHz) at 55°C for 30 min. The suspension was sonicated again for 2 h at 30 °C and left to rest overnight at RT to stabilize the interaction. GNRs were then centrifuged at 5600g for 10 min and suspended in an aqueous solution. The GNR-LA complexes were kept at 4°C in the dark. Next, the modified GNRs were re-dispersed in a 10 mM phosphate buffer containing 16 mM EDAC and 4 mM sulfo-NHS (30 min in an ice-bath under sonication). After another centrifugation step, GNR-LA suspensions were blocked with poly (ethylene glycol)-thiolate (5kD mPEG-SH, 10-4 mM, from Nanocs) for 10 min in an ice bath, under stirring (Supplemental Fig.1a-b).

TEM images of the nanosensors were obtained on a 120 kV FEI Technai G2-12 (Spirit BioTwin, USA) microscope (Supplemental Fig.1c). Samples were directly dripped onto a holey carbon film supported on a copper grid (400 mesh) (Pelco®, USA) without any further processing. Zeta potential was obtained with a Zetasizer Nano ZS90 analyzer from Malvern at an angle of 173° at RT, as previously described(9-11). Nanoparticle’s size and zeta potential were measured simultaneously three times and in triplicate (Supplemental Fig.1d-e).

Samples’ labeling and measurements were previously described^3,4^. Briefly, GNR-S were incubated with 1mg/mL of anti-S protein polyclonal antibody (Rhea Biotech, Brazil). Next, the samples were labeled with goat anti-rabbit IgG-CFL 488, 100µg/mL λex: 488nm, λem: 520nm (Santa Cruz Biotechnology, USA) (Supplemental Fig1.f). For GNR-N, samples were incubated with 10µg of a polyclonal antibody anti-protein N for 60 min under axial shaking. Unbound antibodies were blocked with BSA (1%) for 30 min. Next, samples were labeled with Alexa Fluor® 546 goat anti-rabbit IgG H & L, 5µg/mL λex: 540nm, λem: 585nm (Invitrogen, USA) and incubated under continuous shaking in the dark and at RT for 30 min (Supplemental Fig.1g). Samples were measured using the spectrophotometer Varioskan Flash spectral scanning multimode reader (Thermo Scientific, USA).

To determine the concentration of antibodies in the LSPR-nanosensor, an antibody curve was carried out ranging from 0.25µg to 10µg for anti-S and anti-N proteins, respectively (Supplemental Fig.1h-l). The concentration chosen was 0.5 µg for both GNR-S and GNR-N nanosensors. Afterward, the nanosensors were exposed to purified S- and N-proteins, ranging from 10 µg to 10fg, and their binding affinity was assessed by UV-Vis spectroscopy (Thermo Scientific, USA).

Finally, the GNR-S and GNR-N nanosensors were exposed to macerated testes samples (obtained from the 11 patients’ with COVID-19), diluted in PBS-1X, for 30min at 4°C under sonication, and their respective LSPR were measured using optical plates (Costar, USA), by UV-Vis spectroscopy (Thermo Scientific, USA). Spectra analyses were performed using OriginPro version 9.0. We normalized and smoothed the curves and measured the biosensing event through the X-axis intercept of the derivative of the Gaussian peak for each patient. We compared the COVID-19 patient curves to the GNR and negative control patient curves (Supplemental Fig.1 m-q). We focused on the redshift that occurred at point zero (derivative axis) of the longitudinal peak. In this analysis, 5nm or below shifts were not considered significant.

**Fig. 1.**
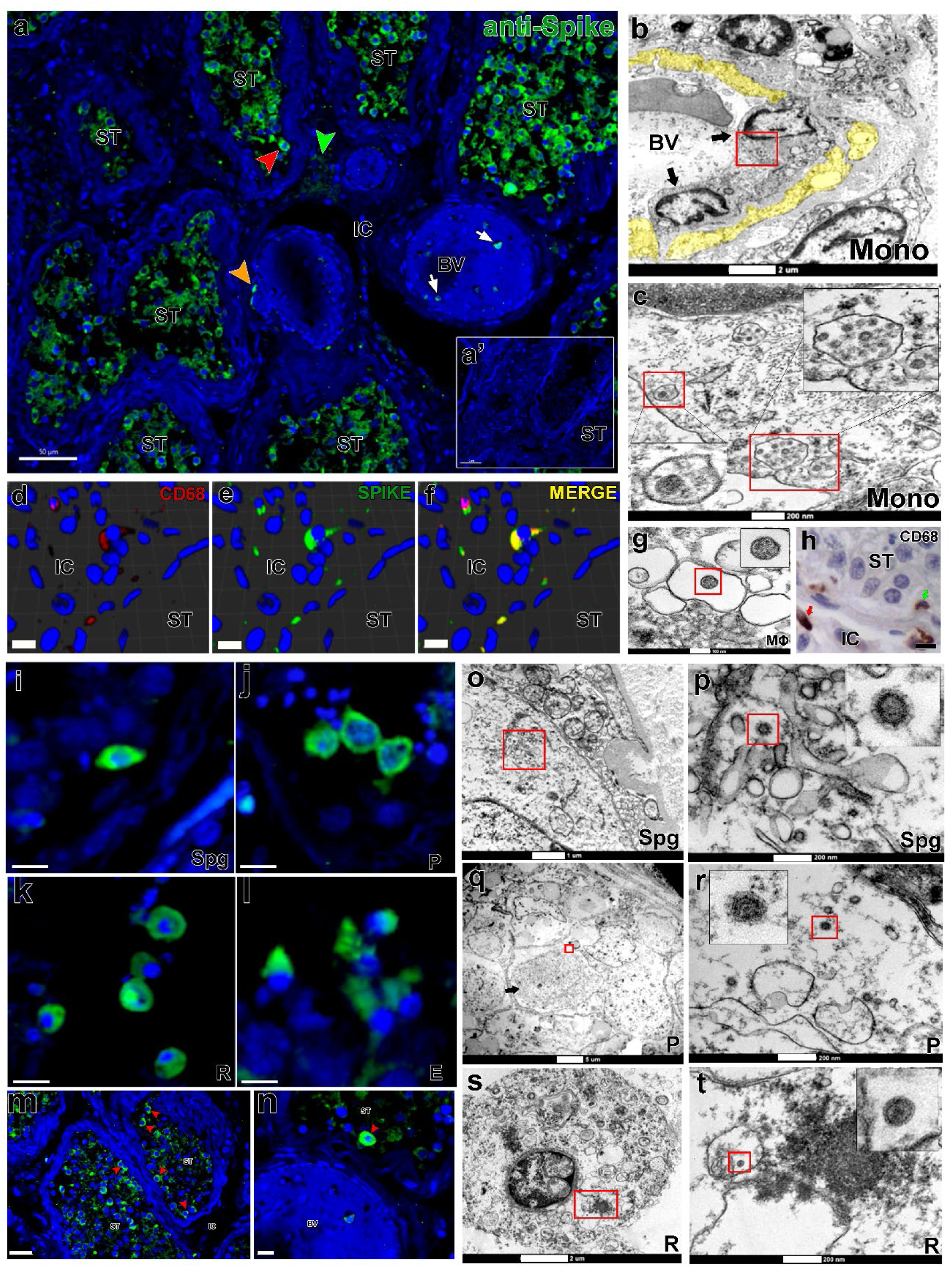
SARS-CoV-2 infection in monocytes, macrophages and germ cells. a) immunofluorescence for S-protein in testicular parenchyma showing positive germ cells (red arrowhead), monocytes (white arrow), macrophages (orange arrowhead) and Leydig cells (green arrowhead); a’) negative control omitting the primary antibody (Scale bars = a: 50µm; a’: 40µm). b-c) TEM images showing infected infiltrative monocytes (Mono, arrows); Yellow = disrupted cytoplasm of endothelial cells; Red boxes = cytoplasmic areas replenished with viral particles; Inserts = high magnification of viral particles (Scale bars = g: 2µm; h: 200nm). d-f) 3D reconstruction of a double immunofluorescence for CD68 (w, red color) and S-protein (x, green color), depicting an infected macrophage (y, merge – yellow color) (Scale bars = 10µm). g) TEM image of an infected macrophage (m) showing a viral particle in its cytoplasm (insert). h) Presence of macrophages (arrows) inside the seminiferous tubule (green arrow) and in the interstitial area (red arrow) (Scale bar = 10µm). i-l) different types of infected germ cells: i: Spermatogonial cell (Spg); j: Pachytene spermatocytes (P); k: Round spermatids (R); l: Elongated spermatids (E) (Scale bars = 15µm). m-n) immunofluorescence against the S-protein showing that spermatogonial cells (red arrowheads) are more intensely labeled than other germ cells. (Scale bars = a: 30µm; b: 10 µm). o-t) TEM images showing viral particles (in low and high magnification) in germ cells, as follows: o-p) Spermatogonial cell (Spg) (Scale bars = a: 1µm; b: 200nm); q-r) Pachytene spermatocytes (P, arrow) (Scale bars = c: 5µm; d: 200nm); s-t) Round spermatids (R) (Scale bars = e: 2µm; f: 200nm). Immunofluorescence images in the testis of patient #8. TEM images in testes from patients #1, #7 and #8. Blue = dapi counterstaining; green staining = IgG-CFL 488; red staining = IgG-546. ST = seminiferous tubule cross-section; IC = intertubular compartiment; BV = blood vessel.

#### Immunofluorescence against Spike protein

Immunofluorescence was performed using a validated primary anti-S protein antibody(12) (Rhea Biotech; IM-0828) to detect and corroborate the viral presence in the testicular parenchyma (Supplemental Table 2). Reactions were visualized using Alexa-488 (1:100 dilution; Thermo Fisher Scientific, USA) conjugated secondary antibody, and images were acquired using a Nikon Eclipse Ti fluorescence microscope. As controls, we performed three different assays: 1) we used the testes of Controls (prostate cancer patients); 2) we omitted the primary antibody in testes of COVID-19 patients; and 3) we performed a negative control antigen, incubating the primary antibody with purified Spike protein (1:10; donated by CT-Vacinas-UFMG) before the IF assay (Supplemental Fig.2m-p).

### SARS-CoV-2 ACTIVITY AND REPLICATION

#### *in vitro* isolation of SARS-CoV-2

Viral isolation was performed to verify if the virus detected in the testes’ samples were infective. In a biosafety level 3 (BSL-3) laboratory, a sample of the COVID-19 patient #1 was macerated using the L-Beader 6 tissue disruptor (Loccus, Brazil) and magnetic beads. We chose this patient because he presented the earliest CT for SARS-CoV-2 gene N in the RT-qPCR (with specific viral primers). The disrupted sample was resuspended in 500 µl of PBS 1x, and 200 µL was used for infection *in vitro*. Vero CCL-81 cells were seeded in a T25 flask until complete confluence was achieved. Culture media was discarded, and cells were incubated with 200 µL of the disrupted sample for 1 h, at 37°C and 5% CO2, with periodic homogenization.

After incubation, 5 mL of fresh DMEM was added, and cells were maintained (at 37°C and 5% CO2) for 72 h or until cytopathic effects. In parallel, the same process was performed on a 24-well plate. The results were compared with a Mock-infection (Control) in both cases. After this period, supernatant from the T25 flask was collected for viral genome detection by RT-qPCR. Additionally, cells from the 24-well plate were incubated with paraformaldehyde 4% for 20 min (Synth, Brazil) to perform the immunocytochemistry analyses.

#### Viral detection in Vero cell culture

To confirm the isolation of SARS-CoV-2, we extracted the total RNA from the supernatant using a TRIzol extraction protocol. In a 1.5mL tube, 300 µL of supernatant was homogenized with 200 µL of TRIzol (Thermo Fisher Scientific, USA) and incubated for 20 min to guarantee viral inactivation. RNA separation, precipitation, and elution were performed by a chloroform-isopropanol protocol, according to the TRIzol manufacturer’s instructions. According to the kit’s instructions, total RNA was submitted to RT-qPCR using the AllplexTM 2019-nCoV Assay (Seegene, Brazil). Cycling steps were performed in a QuantStudio™ 3 Real-Time PCR System (Applied Biosystems, USA).

#### Immunohistochemistry in Vero cells

To confirm viral isolation from the patients’ testes, we performed an immunohistochemistry assay using the anti-S commercial antibody (Rhea Biotech, Brazil). After fixation, the plates were washed with PBS 1X and blocked with a PBS solution containing 3% FBS for 15 min. Then, the cells were incubated overnight with the anti-S antibody (dilution 1:500) at RT.

The cells were also incubated with an anti-rabbit IgG antibody conjugated to horseradish peroxidase (Promega, USA), diluted at 1:2500 at RT for 60 min. Immunoassay was revealed using the KPL TrueBlue Peroxidase Substrate (SeraCare, USA) for 10 min at RT, under gentle stir.

### HISTOMORPHOMETRIC ANALYSIS

All testicular slides were scanned using the Panoramic MIDI II slide scanner (3DHISTECH, Hungary). The histomorphometric analyses were performed using the CaseViewer software (3DHISTECH, Hungary) and the Image J v.1.45s software (Image Processing and Analysis, in Java, USA).

#### Seminiferous epithelium cell composition

To describe the seminiferous epithelium integrity, at least 50 seminiferous tubules cross-sections were evaluated and classified according to the presence of the most differentiated germ cell type. The cross-sections were then classified as 1) containing all germ cell layers; 2) containing spermatogonia and spermatocytes; 3) containing only spermatogonia, and 4) degenerating/Sertoli cell-only seminiferous tubules. After this blinded analysis, to understand the evolution of testicular pathogeny, we examined the clinical data of COVID-19 patients. These patients were classified into three phases according to the severity of the damages inflicted to the seminiferous epithelium. The results are presented as the percentage of seminiferous tubules in each category.

#### Seminiferous tubule measurements

Seminiferous tubules were analyzed using computer-assisted image analysis of 30 randomly chosen seminiferous tubules cross-sections per donor. To determine the seminiferous tubule diameter and tunica propria width, the measurements were taken at 400x magnification, and the results were expressed in µm.

#### Leydig cell volume density

The volume densities (%) of testicular tissue components were obtained after counting 7200 points over testis parenchyma. The intersections that coincided LCs were counted in 15 randomly chosen fields by horizontal scanning of the histological sections at 200x magnification(13).

#### Hemorrhagic scores

As red cell bleeding was a common finding, this pathology was measured in four scores, as follows:

1. Patients who presented many red blood cells inside the seminiferous tubule lumen and intertubular compartment (score 3);
2. Patients with vast areas of red blood cells bleeding in the intertubular compartment (score 2);
3. Patients with small areas of red blood cell bleeding in the intertubular compartment (score 1);
4. Patients without red blood cell bleeding (score 0).

### HISTOCHEMISTRY TECHNIQUES

#### Mast cell counts

Toluidine-blue staining was used to determine the number of mast cells. We investigated 15 testicular fields (20x magnification), and the cells were quantified per patient.

#### PAS staining

A Schiffs kit (Sigma-Aldrich, USA) was used for PAS staining, as per the manufacturer’s protocol. In brief, the sections were pre-treated with periodic acid for 5 min at RT, slowly rinsed in distilled water, and then stained with Schiffs solution for 10 min at RT in the dark. The nuclei were stained with hematoxylin for 5 min at RT, followed by six dips in 1% hydrochloric alcohol. After dehydration with 70, 90, and 100% graded alcohol, the sections were immersed twice in xylene for 10 min each. Then, the slides were mounted with a coverslip and sealed with Entellan resin (Sigma-Aldrich, USA). Images were captured using light Olympus microscopy (BX-60).

#### Masson’s Trichrome and Picrosirius red

Tissue samples were stained with Masson’s Trichrome and Picrosirius red to assess fibrosis and collagen types I and III. Images of the two techniques were captured in a Spot Insight Color digital camera adapted for Olympus BX-40, using the Spot software version 3.4.5. To determine the area of fibrosis and differentiate the types of collagens, images of three random areas of each patient were obtained at 100x magnification. Images were analyzed with Image J v. 1.53c software (National Institutes of Health, USA) using the Color Deconvolution tool, getting the average of the three areas evaluated(14).

### IMMUNOSTAINING

For immunostaining, deparaffinized sections (5 μm thick) were dehydrated and submitted to heat-induced antigenic recovery (water bath) with buffered sodium citrate (pH 6.0) at 90°C for 40 min. Then, the sections were immersed in BSA 3% (in PBS) solution to block non-specific antibody binding and incubated overnight at 4 °C with primary antibodies (Supplemental Table 2). Reactions were visualized using biotin-conjugated secondary antibodies (anti-goat: 1:100 dilution, Abcam, ab6740; anti-mouse: 1:200 dilution, Imuny, IC1M02; anti-rabbit: 1:200 dilution, Abcam, ab6720) combined with Elite ABC Kit (Vector Laboratories, USA). Signal detection was obtained via peroxidase substrate 3,39-diaminobenzidine (DAB; Sigma Aldrich, USA) reaction and counterstaining with Mayer’s hematoxylin (Merck, USA).

To identify targets of SARS-CoV-2 in the testis intertubular compartment, double-immunofluorescence anti-S with anti-CD-68 or anti-Chymase was performed. Reactions were visualized using Alexa-488 (1:100 dilution), Alexa-546 (1:200 dilution), and Alexa-633 (1:200 dilution), all from Thermo Fisher Scientific, USA, and conjugated secondary antibody. For ACE2 evaluation, we used anti-ACE2 (Proteintech, USA) and Alexa-594 (1:100 dilution) from Jackson Immunoresearch, USA. Images were acquired using the Nikon Eclipse Ti fluorescence microscope.

To analyze immunopositive macrophages and T lymphocytes, CD68 and CD3 positive cells were quantified by counting 10 testicular fields at 400x magnification. Only cells with visible nuclei, brown cytoplasm, and morphology compatible with the evaluated cells were counted(15). Microvessel density analysis was also performed in 10 fields at 400x magnification, and only CD31 immunopositive structures with or without lumen were counted (vessels containing muscle walls were not counted).

### TRANSMISSION ELECTRON MICROSCOPY

Testes fragments were fixed by immersion in 4% glutaraldehyde (EMS, USA). Smaller pieces (1-2 mm thickness) were obtained and postfixed in reduced osmium (1% OsO4 and 1.5% potassium ferrocyanide in distilled water) for 90 min, dehydrated in ethanol, and embedded in Araldite epoxy resin. Ultrathin sections (60 nm thick) were obtained using a diamond knife on a Leica EM UC6 ultramicrotome (Leica Microsystems) and mounted on 200 mesh copper grids (Ted Pella). The ultrathin sections were stained with lead citrate (Merck, USA) and analyzed using a transmission electron microscope (Tecnai G2-12 Thermo Fisher Scientific/FEI, USA).

### ENZYMATIC AND HORMONE MEASUREMENTS

The enzymatic activity of Myeloperoxidase (MPO) and N-Acetyl-BD-glucosaminidase (NAG) and the total concentration of testosterone and angiotensin II were determined in human testis homogenates. For this purpose, 50 mg of snap-frozen testis from deceased COVID-19 patients and Controls was homogenized in 450 µL cold PBS supplemented with a protease inhibitor cocktail (Cat n° S8830, Sigma-Aldrich). After three freeze/thaw cycles in a liquid nitrogen/water bath (37°C), the samples were centrifugated (14.000 g, 10 min, 4°C), and the supernatants were collected.

For MPO and NAG measurements, 100 µL of tissue homogenates were mixed 1:1 in MPO buffer assay (0,1M Na3PO4, 0.1% [w/v] HETAB, pH 5.0) or NAG buffer assay (0,2M citric acid, 0,2M Na2HPO4, pH 4.5), respectively, just prior the freeze/thaw step. The activity of MPO and NAG, which is an indirect estimation of the abundance of neutrophils and macrophages, was measured in a colorimetric enzymatic assay. Intratesticular testosterone levels were determined in testis homogenates using a chemiluminometric immunoassay run on the Atellica IM Analyzer (Siemens Healthcare Diagnostics). The concentration of Angiotensin II was measured by ELISA, according to the procedures supplied by the manufacturer (MyBioSource, San Diego, CA, USA). The kit applied the sandwich ELISA technique. The sensitivity of the assay was 12 pg/mL.

### TESTICULAR GENE EXPRESSIONS

Total RNA was isolated from testes using AurumTM Total RNA MiniKit® (BioRad, USA). A Nanodrop spectrophotometer (Thermo Fischer, USA) was used to measure the quantity and integrity of total RNA. RNA (2 μg/sample) was reverse transcribed using the iScript cDNA Synthesis Kit (BioRad, USA). cDNAs (10 ng) were amplified by qPCR with iTaq Universal SYBR Green Supermix (BioRad, USA) in Rotor-Gene Q (Qiagen, USA). The primer sequences used can be found in Supplemental Table 2. Relative levels of expression were determined by normalization to RPL19 e HPRT1 using the ΔΔCT method. The testicular gene expressions were displayed as Heat maps (Fig.5, Fig.5 and Fig.6) and individual graphs (Supplemental Fig.6 and Supplemental Fig.7).

### STATISTICAL ANALYSES

Demographics and clinical characteristics were presented using descriptive statistics: mean and standard deviation (SD) for normally distributed continuous data; median and interquartile range (IQR) for non-normally distributed continuous data; and proportions and frequencies for categorical data. The presence and the strength of a linear relationship between demographics and clinical characteristics and the damage to the testicles were analyzed using Spearman correlation.

All quantitative data were tested for normality and homoscedasticity of the variances following Kolmogorov–Smirnov (Dallal–Wilkinson–Lilliefor) and Bartlett tests. Data from the fluorometry assay analyses were evaluated by one-way ANOVA for comparisons within groups, followed by Newman-Keuls (normal distribution). Histomorphometric and gene expression data were analyzed by unpaired Student’s t-test, comparing COVID-19 groups to Controls and COVID-19-P1 to COVID-19-P2 (COVID-19-P3 was not considered for statistics). The data obtained were represented as the mean ± SEM and geometric mean ± SD. Graphs and statistical analyses were conducted using GraphPad PRISM v6.0 (GraphPad Software, Inc). Differences were considered statistically significant at p< 0.05.

## RESULTS AND DISCUSSION

### Hypertension, diabetes, and obesity are the main comorbidities

All 11 patients studied herein were admitted to the ICU in in Belo Horizonte (Brazil) hospitals due to severe pulmonary symptoms. Mean age was 63.9 ± 13.11 years (range 46 to 8) and the mean body mass index was 32.18 Kg/m2 ± 6.04 (range 25.95 – 44.46). All patients had children, except for patient #9.Table 1 summarizes the patients’ clinical characteristics. None of the patients presented scrotal symptoms or complaints during their hospital stay, nor their clinical history revealed previous testicular disorders. The mean disease duration (from onset to death) was 23.73 ± 8.24 days (range 13–38), while the meantime of ICU admission to death was 15.81 ± 6.19 days (range 8–25). All patients presented fever during hospitalization, with six of them for longer than 24 hours. However, we did not evidence a strong correlation between fever and the development of testicular pathogenesis (Spearman’s = -0.195; p= 0.57). Recent data indicate that fever is not correlated with the sperm quality alteration after SARS-CoV-2 infection(16). The most prevalent comorbidities were systemic arterial hypertension (nine cases), diabetes mellitus (six cases), and obesity (six cases).

### SARS-CoV-2 reliable detection in testes

We first tested the testicular tissue using a conventional RT-qPCR protocol for SARS-CoV-2 and only patient #8 was positively detected (cycle threshold - CT=38). To improve detection and reduce interference of intrinsic tissue factors, we performed a cDNA synthesis using SARS-CoV-2 specific viral primers. The RT-qPCR revealed the virus presence in 10 of 11 patients (Table 2). Previous studies presented conflicting results regarding the detection of SARS-CoV-2 RNA in the testes(2-5).

**Table 2.** SARS-CoV-2 detection in testicular parenchyma.

|  |  | Genetic evaluations |  |  |  | Protein evaluations |  |  |
| --- | --- | --- | --- | --- | --- | --- | --- | --- |
|  |  | SARS-CovV-2<br>E gene<br>RT-qPCR<br>(routinary<br>diagnosis)<br>(CT) | RNP<br>control<br>RT-qPCR<br>(CT) | SARS-CoV-2 (N1)<br>RT-qPCR<br>(using viral<br>primers)<br>(CT) | β-actin<br>control<br>RT-qPCR<br>(CT) | LSPR-nanosensor<br>N shift<br>(nm) | LSPR-nanosensor<br>S shift<br>(nm) | IF using<br>anti-S<br>protein<br>(- or +) |
|  | **PATIENTS** |  |  |  |  |  |  |  |
|  | **CTRL #1** | - | 20 | - | 18.55 | 0 (-) | 0 (-) | - |
|  | **CV-19 #1** | - | 23 | 32.78 (+) | 24.59 | 31.5 (+) | 51 (+) | + |
|  | **CV-19 #2** | - | 27 | 36.99 (+) | 18.50 | 13.2 (+) | 18.9 (+) | + |
|  | **CV-19 #3** | - | 22 | 36.34 (+) | 6.36 | 1.2 (-) | 17.5 (+) | + |
|  | **CV-19 #4** | - | 20 | 35.96 (+) | 19.25 | 11.2 (+) | 17 (+) | + |
|  | **CV-19 #5** | - | 20 | 36.14 (+) | 20.03 | 9.9 (+) | 20.9 (+) | + |
|  | **CV-19 #6** | - | 21 | 36.22 (+) | 10.30 | 13.2 (+) | 15 (+) | + |
|  | **CV-19 #7** | - | 20 | 35.37 (+) | 17.67 | 4.5 (-) | 12.4 (+) | + |
|  | **CV-19 #8** | 38 (+) | 20 | 35.95 (+) | 19.34 | 17.5 (+) | 17 (+) | + |
|  | **CV-19 #9** | - | 21 | - | 19.81 | 12.9 (+) | 17.5 (+) | + |
|  | **CV-19 #10** | - | 23 | 34.03 (+) | 20.71 | 7.2 (+) | 13.6 (+) | + |
|  | **CV-19 #11** | - | 23 | 35.65 (+) | 16.27 | 8.7 (+) | 16.5 (+) | + |

Symbols = CV-19: COVID-19 patient; RNP: RNaseP gene; CT: cycle threshold; LSPR: localized surface plasmon resonance; IF:
immunofluorescence; (+) detection of viral content; (-) non-detection of viral content. Positivity in the RT-qPCR is detected when the CT is lower
than 40, and in the LSPR nanosensor is when the shift is higher than 5 nm.

To confirm the genetic data, we used a nano-designed sensor (which employs localized surface plasmon resonance - LSPR)(11) to detect the SARS-CoV-2 Spike (S)- protein and the nucleocapsid (N)-protein (Supplemental Fig.1a-q). S-protein was found in all, while N-protein was observed in nine patients (Table 2 and Supplemental Fig.1m-q). Prominent S-protein immunolabeling was evidenced in testes of all COVID-19 patients (Supplemental Fig.2), especially in patient #8 (positively detected by all methodologies). These findings suggest that the SARS-CoV-2 tropism for testes is higher than previously thought and that conventional RT-qPCR protocol may only detect infected testes with a higher viral load. Our data suggest that more sensitive techniques are required for the reliable detection of SARS-CoV-2 (even in a low viral titer) in testes.

**Fig. 2.**
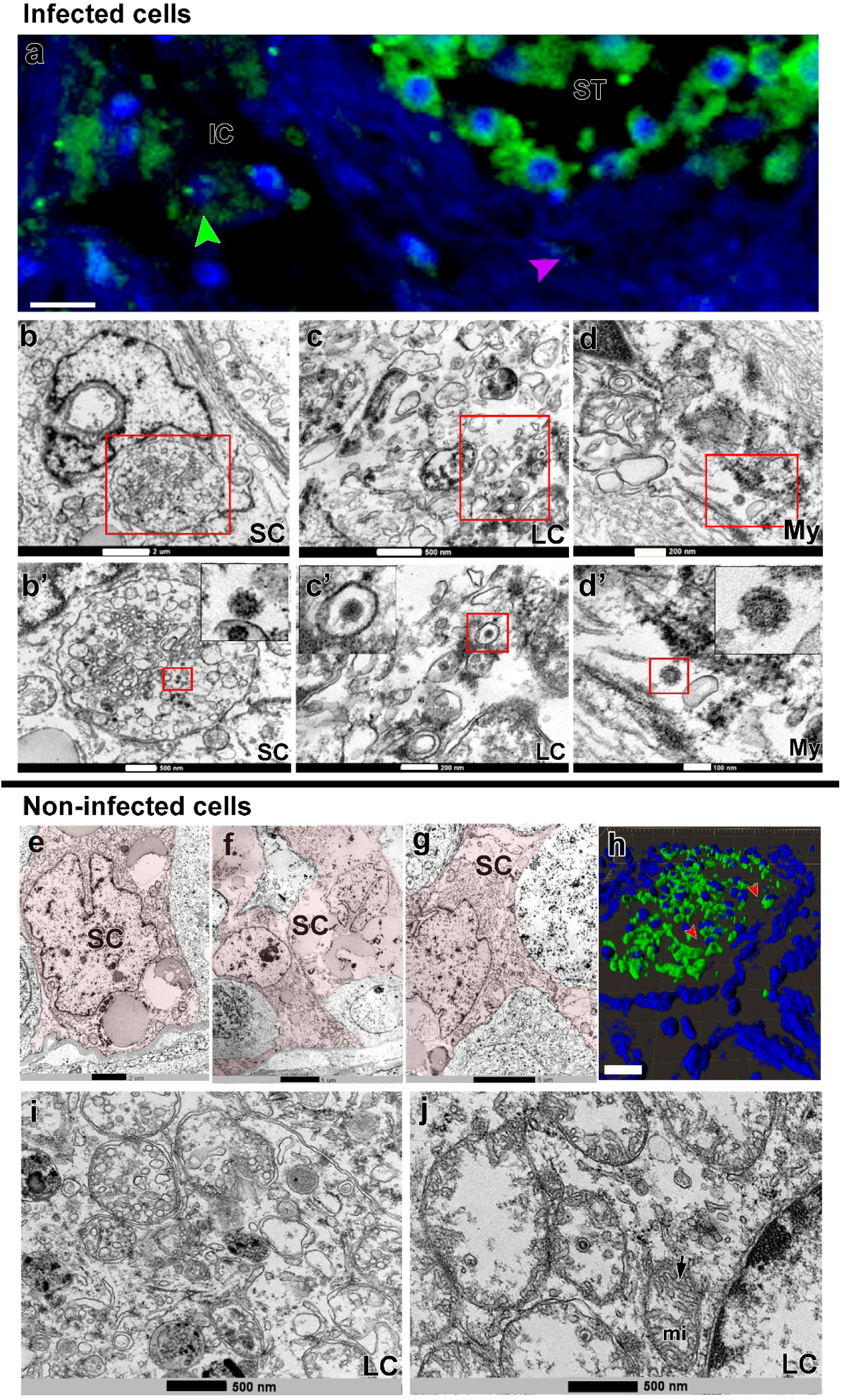
SARS-CoV-2 infection in Sertoli, Leydig and peritubular myoid cells. a) Immunofluorescence against S-protein evidencing weak labeling in peritubular myoid (pink arrowhead) and Leydig cells (green arrowhead) (Scale bar =15µm). b-d’) TEM images showing viral particles (in low and high magnification) in Sertoli cell (SC) (Scale bars = b: 2µm; b’: 500nm); Leydig cell (LC) (Scale bars = c: 500nm; c’: 200nm and peritubular myoid cell (My) (Scale bars = d: 200nm; d’: 100nm). e-g) TEM images of non-infected Sertoli cells (SC, pink) (Scale bars = e: 2µm f-g: 5µ). h) 3D reconstruction of a seminiferous tubule cross-section showing non-labeled areas surrounding germ cells (red arrowheads) (Scale bar = 40µm). i-j) high magnification of non-infected Leydig cells. Arrow = tubular crest of a mitochondria (mi) (Scale bars = 500nm). Immunofluorescence images in the testis of patient #8. TEM images in testes from patients #1, #7 and #8.

### Main infected cells in testis parenchyma

Several infected monocytes/macrophages (CD68+) were detected surrounding blood vessels and migrating to the parenchyma (IF and TEM data), suggesting that these cells might be delivering SARS-CoV-2 to testis (Fig.1a-c, arrows), contributing to infection of testicular cells. Infected macrophages were confirmed through double immunofluorescence and TEM (Fig.1d-g). Monocytes/macrophages were also found inside the tubular compartment, indicating a possible route for viral spreading inside the seminiferous tubules (Fig.1h). Conversely, chymase positive mast cells were not labeled for S-protein (Supplemental Fig.3a-b).

Most S-protein labeling was identified inside the seminiferous tubules, mainly in germ cells (Fig.1 i-m). While we show germ cells positive for the S-protein, other authors showed positivity for the N-protein(4). In some areas, spermatogonial cells displayed intense S-protein labeling compared to spermatocytes and spermatids (Fig.1n, red arrowhead). Nevertheless, SARS-CoV-2 S-protein was detected in spermatogonia, spermatocytes and spermatids (Fig.1o-t). Patients who died up to 20 days after intensive care unit (ICU) admission presented the highest mean fluorescence index in the testis (Supplemental Fig.2a). Germ cells infected with SARS-CoV-2 increase the concerns of potential sexual transmission, reinforced by the detection of SARS-CoV-2 RNA in semen of patients who suffered the severe form of COVID-19(17).

Some Sertoli, Leydig (highly express ACE2(18)), and peritubular myoid cells also presented viral particles in their cytoplasm (TEM data), albeit with lower S-protein immunolabelling intensity (Fig. 2a-c’, arrowheads and insets). However, many of these cells did not show obvious viral particles in the cytoplasm (TEM data) (Fig. 2e-j).

### Viral replication and viability

TEM analyses identified a viral replication factory in macrophages, which are known to express ACE2 and TMPRSS2(19), and in spermatogonial cells, which highly express TMPRSS2(18) (Fig.3a-f). In both cell types, we observed SARS-CoV-2 replication complexes with the formation of convoluted membranes (replication membranous webs, RMW) containing double-membrane vesicles (DMV) and Endoplasmic Reticulum Golgi Intermediate Complex (ERGIC) showing new virions (Fig.3c-f). These morphological features are in accordance with SARS-CoV-2 replication in the cell cytoplasm(20).

**Fig. 3.**
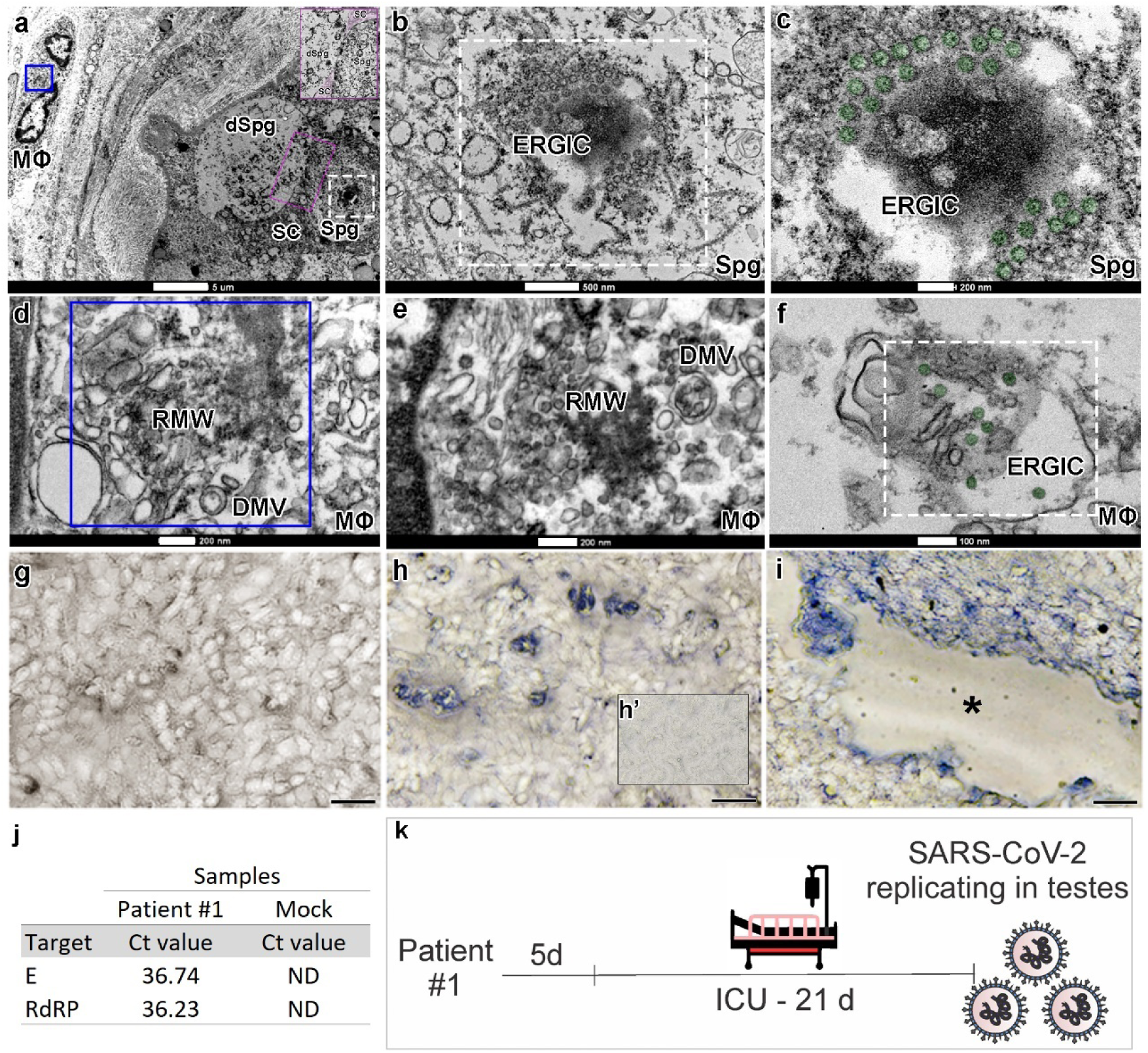
Infective SARS-CoV-2 particles formation and replication activity in Vero cells. a-f) TEM images of testicular parenchyma (Scale bars = a: 5µm; b: 500nm; c: 200nm; d: 200nm; e: 200nm; f:100nm). a) altered seminiferous tubule cross-section, depicting a macrophage in tunica propria containing a Replication Membranous Web (RMW; blue box) and a spermatogonial cell displaying an Endoplasmic Reticulum Golgi Intermediate Complex (ERGIC, white box). The pink box indicates a spermatogonial cell clone surrounded by the cytoplasm of a Sertoli cell (labeled in pink). b-c) higher magnifications of spermatogonial cells showing new virions (labeled in green) inside the ERGIC. d) higher magnification of the RMW from the macrophage depicted in “figure a”, evidencing the presence of double-membrane vesicles (DMV). e-f) images from an interstitial macrophage presenting RMW, DMV, and ERGIC filled with new virions (labeled in green). g-i) infectivity of SARS-CoV-2 was evaluated in VERO CCL-81 cells (Scale bars = 30µm). g) cell confluence of VERO CCL-81 cultures prior exposition to testicular macerate. h) S-protein immunostaining showing infected VERO CCL-81 (blue labeling: KPL TrueBlue Peroxidase Substrate). h’) negative control omitting the primary antibody. i) cytopathic effect in VERO CCL-81 cell culture (empty area, *) after being exposed to testicular homogenate for 72 hours. j) viral load in the VERO CCL-81 cell culture supernatant was measured using RT-qPCR. Mock = control group; ND = non-detected. k) schematic drawing indicating that SARS-CoV-2 remains active and replicating in gonads 26 days after the onset of clinical symptoms.

The testicular immune privilege prevents the autoimmune attack of haploid germ cells, but it also allows viruses to escape immunosurveillance(21). The “macrophage paradox”, described in SARS-CoV-2-induced lung damages(19) may also apply to the testes. Thus, although macrophages combat viral infections, they can act as Trojan horses(19), facilitating viral entrance and replication in testis. While it is possible that the testis can be infected by direct invasion caused by viremia, our results point that infected monocytes/macrophages migrating (e.g., from lungs) may also be actively transporting the virus into the testes(22). Identifying viral access to the testis and the local of replication are relevant because testicular immune tolerance may hamper the viral clearance from the human body, like what is observed for other viruses(21).

In a BSL-3 lab, we exposed VERO CCL-81 cell cultures to testicular homogenates from an infected patient and found that SARS-CoV-2 was infective and able to replicate in these cells. This data was confirmed by immunohistochemistry, the cytopathic effect in the infected VERO cell culture and conventional RT-qPCR (Fig.3g-j), indicating that we were not observing just fragments of viral particles.

Previous research indicated that SARS-CoV-2 negativity in RT-qPCR tests (at least in two consecutive assays) usually occurs between 6-12 days from the time of onset of symptoms(23). However, according to our data, the virus remains viable for a longer period within the testis. Indeed, SARS-CoV-2 was detected in the testis of patient #1, who died 26 days after symptoms’ onset (Fig.3k). Thus, our findings suggest that the testes may serve as sanctuaries for SARS-CoV-2, maintaining infective viruses for extended periods. Testis environment can even be related to the delayed viral clearance in men compared to women(24). Furthermore, the testes may not be neglected in evaluating the patients’ clinical condition because it is a site of viral replication and consequently a source of viral load.

### Clinical data and testicular alterations

Since SARS-CoV and SARS-CoV-2 infection may impair male GC development, leading to GC loss(2,25), we categorized the COVID-19 patients according to the architecture and histology of seminiferous tubules to understand the evolution of the testicular pathogeny, emphasizing the preservation of different generations of germ cells. Critically ill patients who maintained elongated spermatids in the seminiferous epithelium were categorized as the first phase (patients #6, #7, #8, #9, and #11). Patients presenting primary spermatocytes as the most advanced germ cell type were categorized as the second phase (patients #1, #3, #4, #5, and #10). Specifically, patient #2 was classified in the third phase because his testis presented few spermatogonia inside the seminiferous tubules (Fig.4a-c). A weak correlation (Spearman’s = -0.33; p=0.30) was observed between the germ cell loss and the patient’s age. Furthermore, we did not observe a strong correlation between patients’ age and the progression of the pathogeny (Spearman’s = 0.33; p= 0.31).

**Fig 4.**
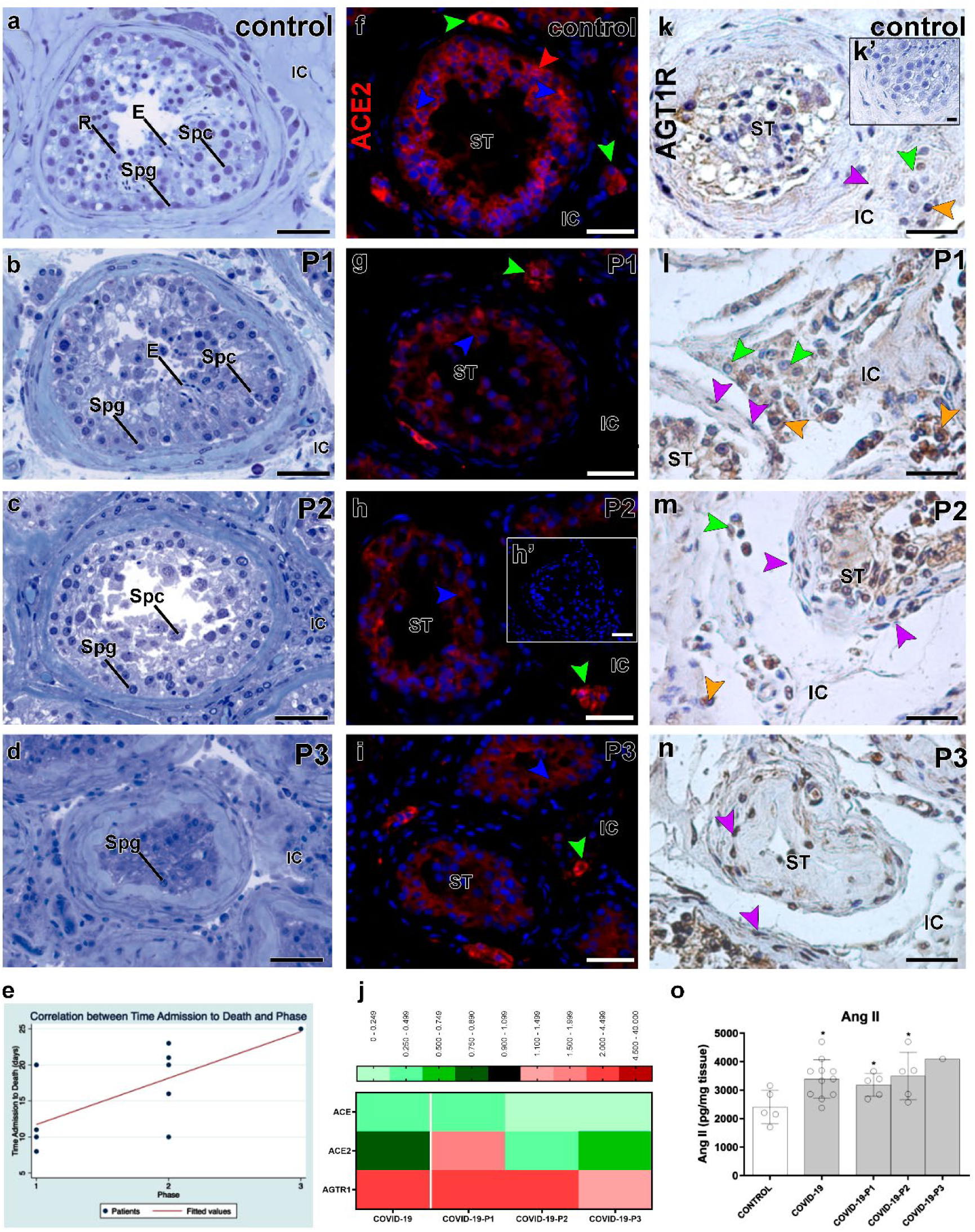
Imbalanced renin-angiotensin system in the testis of COVID-19 critically ill patients. a-d) initial categorization of COVID-19 patients according to the most advanced germ cell in the seminiferous epithelium: phase 1 (P1), phase 2 (P2), and phase 3 (P3). Toluidine-blue counterstaining (Scale bars = 50 µm). a) Control patients showing elongated spermatids (E). b) COVID-19 patients showing elongated spermatids (E), spermatocytes (Spc) and spermatogonial cells (Spg). c) COVID-19 patients displaying spermatocytes (Spc) and spermatogonial cells (Spg). d) COVID-19 patients presenting spermatogonial cells (Spg) only. e) positive correlation between patient’s phases and the time frame from ICU admission to death (Spearman’s rho = 0.6862; p=0.0197). f-i) immunostaining of ACE2 in the testis of Control (f) and COVID-19 patients (g-i) (Scale bars = 50 µm). h’) Negative control, omitting the primary antibody. Red: Alexa-594; Blue: dapi; Green arrowheads: Leydig cell; red arrowheads: spermatogonial cell; blue arrowheads: Sertoli cell. j) heat map of the evaluated genes related to the renin-angiotensin system. The colors compare COVID-19 patients to Controls. Green: lower expression than Controls. Red: Higher expression than Controls. k-n) immunostaining of AGT1R in the testis of Control (k) and COVID-19 patients (l-n). Purple arrowheads: peritubular myoid cells; green arrowheads: Leydig cells; orange arrowhead: macrophages (Scale bars = 50 µm). o) ELISA assay for angiotensin II (t-test; two tailed; infected groups vs control; *p<0.05). P1, P2, P3: phase 1, 2, and 3 of the COVID-19 patients, respectively. ST = seminiferous tubule; IC= intertubular compartment.

**Fig 5.**
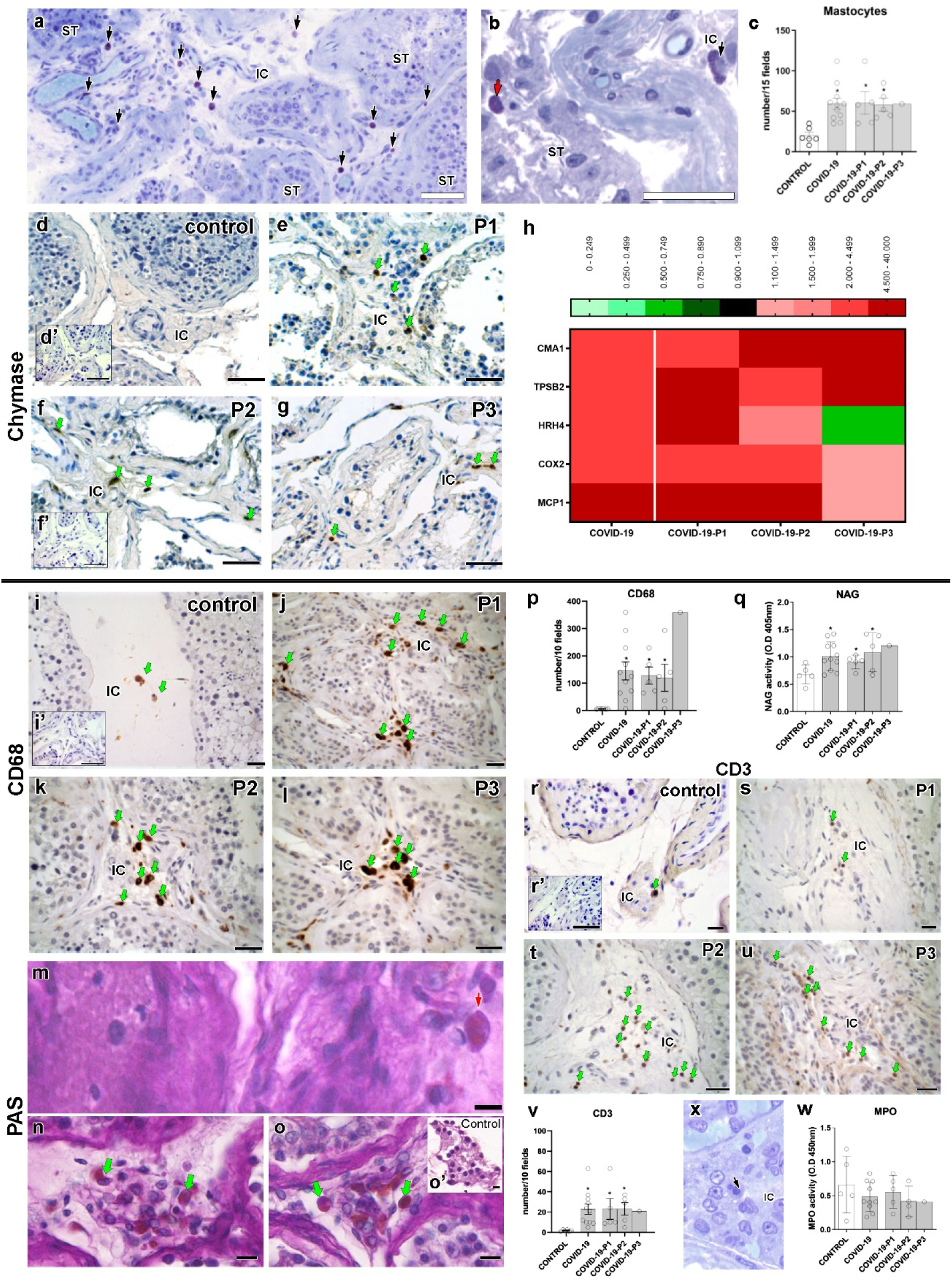
Presence of activated mast cells and macrophages in the testis parenchyma of COVID-19 critically ill patients. a) toluidine blue staining showing an elevated number of mast cells (arrows) in testis parenchyma (Scale bar = 50 µm). b) In some regions, mast cells were identified inside the seminiferous tubules (red arrow). c) Number of mast cells counted in 15 fields of testis parenchyma at 400x magnification (t-test; two tailed; infected groups vs control; *p<0.05). d-g) immunostaining for chymase in testes of Control and COVID-19 patients. Activated mast cells (arrows) were observed in COVID-19 patients only (Scale bars = 50 µm). d’) Negative control (Scale bars = 50 µm). h) heat map of the evaluated genes related to the mast cells and macrophages. The colors compare COVID-19 patients to Controls. Green: lower expression than Controls. Red: Higher expression than Controls. i-l) immunostaining for CD68 in testes of Control and COVID-19 patients, illustrating the high prevalence of this cell in the deceased patients (Scale bars = 50 µm). i’) Negative control (Scale bars = 50 µm). m-o) PAS-positive monocytes (red arrow) and macrophage (green arrow) indicating their activation (Scale bars = 15 µm). o’) PAS-negative cells in the interstitial cells of a Control patient (Scale bar = 10 µm). p) Number of macrophages counted in 10 fields of testis parenchyma at 400x magnification (t-test; two tailed; infected groups vs control; *p<0.05). q) N-acetylglucosaminidase (NAG) assay indicating the activation of macrophages in COVID-19 patients’ testes (t-test; two tailed; control vs infected group; p>0.05). r-v) immunostaining for CD3 and lymphocyte number in testes of Control and COVID-19 patients (t-test; two tailed; infected groups vs control; *p<0.05) (Scale bars = 50 µm). r’) Negative control (Scale bars = 50 µm). x-w) neutrophils and their activity (s: MPO assay) in testis parenchyma (t-test; two tailed; control vs infected group; p>0.05) (Scale bar = 10 µm).

**Fig. 6.**
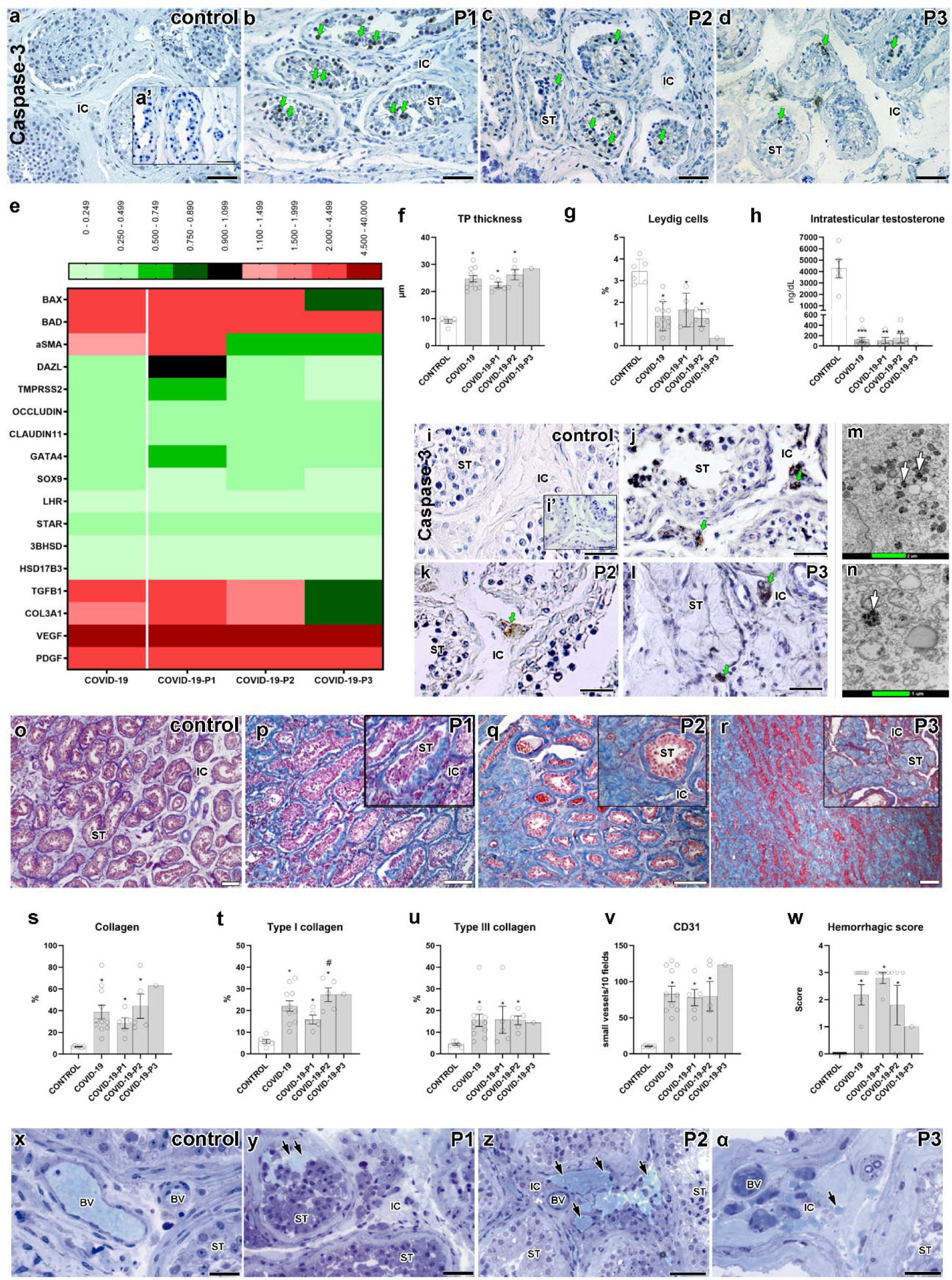
Main testicular dysfunctions following SARS-CoV-2 infection. a-d) immunostaining showing Caspase-3 positive germ cells (green arrows) in testes of Controls and COVID-19 patients (Scale bars = 50 µm). a’) Negative control (Scale bars = 50 µm). e) heat map of critical genes from the tubular and intertubular compartments. The colors compare COVID-19 patients to Controls. Green: lower expression than Controls. Red: Higher expression than Controls. f) measurement of the tunica propria thickness. (t-test; two tailed; infected groups vs control; *p<0.05). g) Leydig cell volume density in testes of Controls and COVID-19 patients (t-test; two tailed; infected groups vs control; *p<0.05). h) measurement of intratesticular testosterone using RIA (t-test; two tailed; infected groups vs control; ***p<0.001; **p<0.01). i-l) immunostaining for caspase 3, evidencing positive labeling in Leydig cells (green arrows) (Scale bars = 50 µm). i’) Negative control (Scale bars = 50 µm). m-n) TEM showing the presence of several electron-dense vacuoles in Leydig cell cytoplasm (white arrows) (Scale bars = m: 2 µm; n: 1 µm). o-r) Trichrome Masson staining in the testis of Controls and COVID-19 patients. In higher magnification, inserts (P1, P2, and P3) show the collagen deposition in the seminiferous tubules (Scale bars = 200 µm). s-u) quantification of total collagen fibers, type I collagen, and type III collagen (t-test; two tailed; infected groups vs control; *p<0.05; COVID-19-P1 vs COVID-19-P2; #p<0.05). v) quantification of newly formed blood vessels (CD31+) (t-test; two tailed; infected groups vs control, *p<0.05). w) quantification of testicular hemorrhage (t-test; two tailed; infected groups vs control, *p<0.05). Testicular hemorrhage was classified as normal (x) and containing high numbers of red blood cells (arrows) in tubular and intertubular compartments (y), a vast number of red blood cells in the intertubular compartment (z), and few red blood cells in the intertubular compartment (α) (Scale bars = 50µm). P1, P2, P3: phase 1, 2, and 3 COVID-19 patients, respectively. BV: blood vessels; IC: intertubular compartment; ST: seminiferous tubule cross-section.

**Fig. 7.**
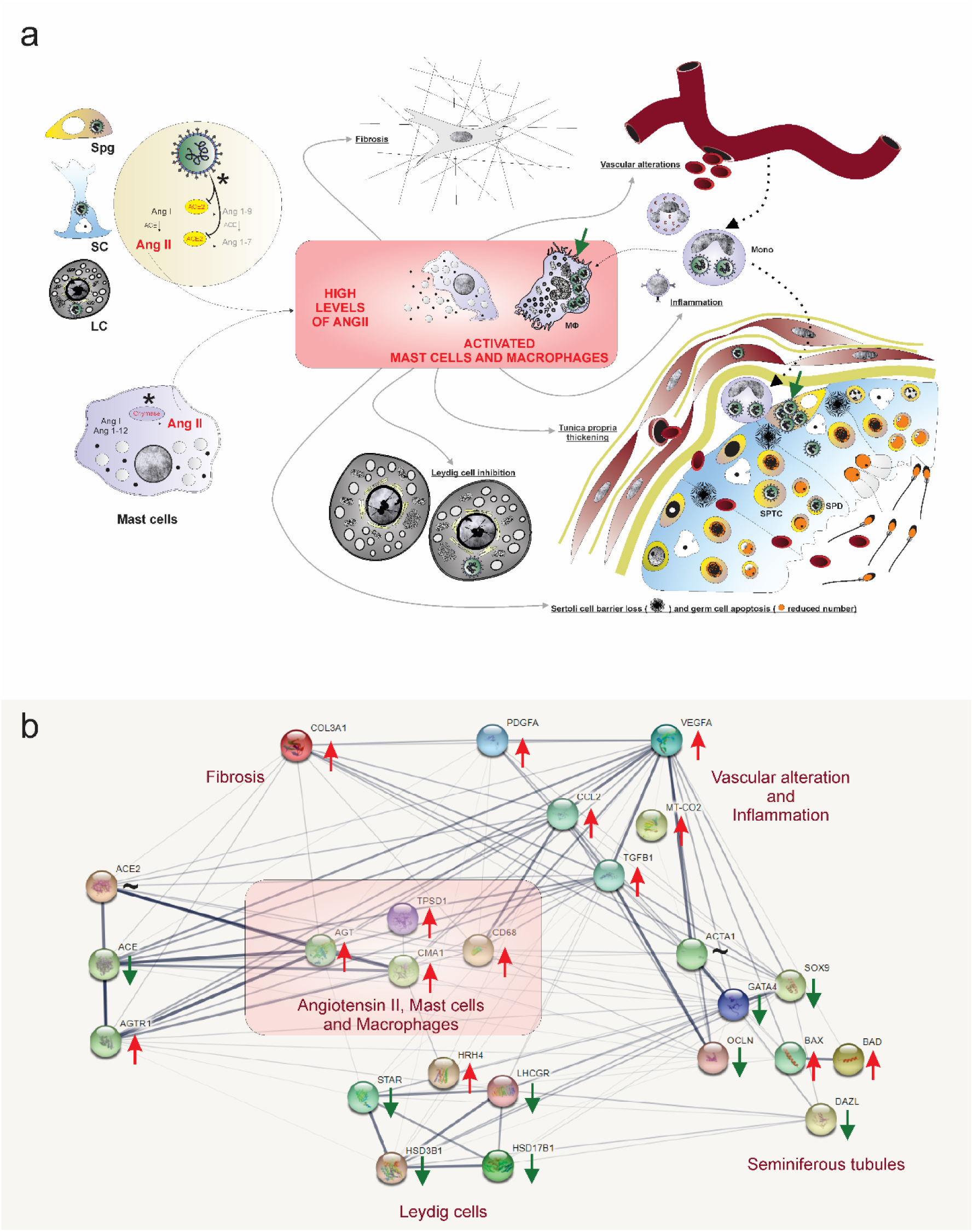
Hypothetical viral and molecular mechanisms of testis infection and damaging by SARS-CoV-2. a) SARS-CoV-2 (green color) was identified in spermatogonial cells (Spg), Sertoli cells (SC), Leydig cells (LC), infiltrative monocytes (Mono), macrophages (MΦ), spermatocytes (sptc), and spermatids (sptd). Note viral factories in macrophages and spermatogonial cells (green arrows). Direct influence of SARS-CoV-2 in testicular cells hampers ACE2 activity, while activation of mast cells (chymase positive) elevates the levels of angiotensin II (a potent pro-inflammatory molecule) (asterisks). Angiogenic and inflammatory factors can induce the infiltration and activation of mast cells. High levels of angiotensin II, activation of mast cells, and inflammatory factors can activate (polarize) macrophages. The testicular phenotype of COVID-19 patients (fibrosis, vascular alteration, inflammation, tunica propria thickening, Sertoli cell barrier loss, germ cell apoptosis, and inhibition of Leydig cells) can be linked to elevated angiotensin II and active mast cells and macrophages. b) genes network related to angiotensin II, activated mast cells, and macrophages (pink box) extracted from STRING (https://string-db.org/). These three elements up-regulate the inflammatory, apoptotic, fibrotic, and vascular genes while down-regulating critical seminiferous tubule and Leydig cell genes. Red arrows: up-regulated genes; Green arrows: down-regulated genes; ∼: genes up-and down-regulated depending on the phase.

Remarkably, there was a positive correlation (Spearman’s = 0.6862; p=0.0197) between the above-described phases and the time from ICU admission to death (Fig.4d).

All patients of Phase I died less than 20 days after ICU admission, with most deceasing well before 20 days. In the second phase, all patients died 20-23 days after ICU admission. Patient #2 (third phase) died after 25 days. Our findings suggest that the more extended severe condition (ICU stay), the lower the number of surviving germ cells. This data raises concerns that patients that survive severe COVID-19 (with an extended stay in ICU) have an increased risk of developing infertility.

### Imbalanced renin-angiotensin system

The interaction of SARS-CoV-2 and ACE2 promotes this enzyme’s internalization, resulting in elevated levels of angiotensin II in the affected tissues(26). We found ACE2 protein in spermatogonial, Leydig, and Sertoli cells of Controls and COVID-19 patient testes (Fig.4f). However, testicular cells of COVID-19 patients showed weak ACE2 staining intensity, specially inside the seminiferous tubules (Fig.4g-i). qPCR revealed augmented ACE2 expression in the testis of first phase patients and diminished in patients of the second and third phases (Fig.4j). ACE protein expression reduced, whereas Angiotensin II Receptor Type 1 (AGT1R) increased in all phases (Fig.4j).

COVID-19 patients presented more intensely immunolabeled testicular cells for the AGT1R than Controls, particularly in Leydig cells, macrophages, and peritubular myoid cells (Fig.4k-n). Angiotensin II levels were higher in the testis parenchyma of COVID-19 patients than in Controls (Fig.4o). Although overstimulation of the ANG II/AGT1R pathway underlies the damages observed in several tissues of COVID-19 patients(27), the present study is the first to show a role for this pathway in the testes.

### Mast cell and macrophage activation

Total protein was significantly higher in testes from COVID-19 patients than Controls (Supplemental Fig.3c), indicating chronic inflammation or infections(28). The influx of immune cells can be mediated by angiotensin II, a potent pro-inflammatory molecule(29), and factors secreted by activated macrophages(30) and mast cells(31). It can also stimulate chemokines that promote mast cell infiltration(32) and regulate migration and infiltration of monocytes/macrophages into tissues(33). Our morphological and molecular data suggest that mast cells and macrophages play a critical role in testicular pathogenesis.

The number of mast cells found in the testis of COVID-19 patients was ten times higher than that found in Controls (Fig.5a-c). Immunohistochemistry revealed chymase-positive mast cells, indicating that these cells are highly active(34) in the testes of the COVID-19-affected patients (Fig.5d-g). Interestingly, mast cells were detected next to areas of the testicular parenchyma displaying a high concentration of immune cells, hemorrhagic areas, altered Leydig cells, interstitial fibrotic areas, damaged tunica propria, and thickened tubular basal membrane (Supplemental Fig.3d-m). qPCR data showed that Control testes present minimal expression of chymase (CMA1) and tryptase (TPSB2), whereas all testicular samples from the COVID-19 patients showed increased expression (Fig.5h). In first phase patients, mast cells were highly prevalent in the testes’ intertubular compartment. Second and third phase patients showed these cells inside the tubular and intertubular compartments (Supplemental Fig.3d-m). Interestingly, increased number of tryptase and chymase positive mast cells in testes is associated with testicular inflammation and infertility in men(35).

Testes of COVID-19 patients also presented elevated mRNA levels of the pro-inflammatory factors MCP1 (10x higher) and COX2 (Fig.5h), which are stimulated by Angiotensin II(36,37). MCP1 is particularly unfavorable for testicular pathogenesis because it can attract more infected monocytes (Supplemental Fig.3n-o).

Macrophages (CD68+) numbers were augmented, and these cells were highly active (NAG assay and PAS staining(38)) in infected testis (Fig.5i-q). Activated macrophages are associated with spermatogenesis impairment or chronic orchitis(39). These cells can compromise the immune-privileged testicular environment, secreting pro-inflammatory cytokines and interfering with testicular homeostasis(40). Therefore, our findings suggest a decisive contribution of activated mast cells and macrophages to the pathophysiology of COVID-19 in the human testes, which could eventually induce harmful effects on the reproductive health in severely ill patients.

Lymphocytes (CD3+) were frequently observed in the testes of COVID-19 patients, suggesting a prolonged infection (Fig.5r-v). Neutrophils were also identified in testes of COVID-19 patients (Fig.5x); however, their activity (MPO assay) was not altered when compared to Controls (Fig.5w). These data suggest that macrophages are more active than neutrophils in the testes of COVID-19 patients.

### Effects on the tubular compartment

While the germ cell population and tubular diameter diminished in COVID-19 patients (Supplemental Fig.4a-b), the tunica propria enlarged, reflecting a higher number of peritubular myoid cells and collagen fibers (Supplemental Fig.4c-j). Interestingly, the tunica propria and the basal membrane (PAS+) thickened next to mast cells and macrophages and sometimes presented a corkscrew appearance (Supplemental Fig.4k-n) There were many caspase-3 positive germ cells in the testes of all COVID-19 patients (Fig.6a-d). The rete testis area presented degenerating germ cells (Supplemental Fig.4o), and BAD and BAX levels were augmented in nearly all COVID-19 patients (Fig.6e). DAZL and TMPRSS2 genes, known to be highly expressed in germ cells, diminished as the phases progressed (Fig.6e).

Tight junctions of the seminiferous tubules were also compromised in COVID-19 patients, with reduced levels of occludin and claudin-11 (Fig.6e). Expressions of critical Sertoli cell genes (Sox9 and Gata4) were also reduced (Fig.6e). Moreover, the seminiferous epithelium detached from the thickened tunica propria (Fig.6f and Supplemental Fig.4p-t), indicating downregulation of tubular junctional proteins. These tubular phenotypes can be stimulated by high levels of angiotensin II(41), activated mast cells(42), and activated macrophages(43). Interestingly, proteases and cytokines released by activated mast cells and macrophages can disrupt tight epithelial junctions and deregulate the human blood-testis barrier(44).

### Leydig cells apoptosis and inhibition

There was a gradual volumetric reduction of Leydig cells (Fig. 6g) as the testicular pathogeny developed, in accordance with Yang et a. (2020)(45). Intratesticular testosterone levels measured in the testicular homogenates of COVID-19 patients were lower (∼30 times) than the Controls (Fig. 6h). Caspase-3 immunolabelling revealed that these cells were undergoing apoptosis (Fig.6i-l). Additionally, their morphology was altered, presenting a vacuolated cytoplasm (with a degenerating content) (Fig.6m-n and Supplemental Fig.5a-d).

Reduction in the expression of LHR, STAR, 3BHSD, and 17BHSD in the testis of COVID-19 patients was consistent with the low number of Leydig cells (Fig.6e). The levels of histamine H4 receptor (HRH4), an inhibitor of human steroidogenesis(46), were highly augmented in the affected patients (Fig.5h). This data indicates that local histamine can inhibit steroidogenesis in the testes of COVID-19 patients.

Mast cells producing histamines, high levels of angiotensin II, and pro-inflammatory cytokines produced by macrophages can inhibit steroidogenesis(47). Moreover, low levels of intratesticular testosterone disturb the immune-privileged milieu(48) and compromise spermatogenesis(49).

### Testis fibrosis and vascular alterations

Testicular fibrosis was commonly observed in the testis parenchyma of COVID-19 patients. Collagen fibers increased progressively (Fig.6o-u), showing high amounts of type I and III collagen in testis parenchyma (Fig.6t-u, Supplemental Fig.5e-h). Further, the expression levels of Col3a were highly augmented in the testes of phase I patients (Fig.6e). It is known that Angiotensin II, activated mast cells, and activated macrophages stimulate collagen synthesis by fibroblasts(50-52).

Expressions of the angiogenic factors VEGF and PDGFA and small vessel volumetric proportion (CD31+) increased in the testis of COVID-19 patients (Fig.6e). Immature blood vessels (TEM and Immunohistochemistry data; CD31+ cells) were observed inside the tunica propria, suggesting an angiogenic process (Fig,6v and Supplemental Fig.5i-m). Red blood cells outside the testicular vasculature were frequently observed in the testes of COVID-19 patients (Fig, 6w-α, arrows). First phase patients presented many red blood cells inside the seminiferous tubule lumen and intratesticular rete testis, possibly due to a reduction in tight epithelial junctions (Supplemental Fig.5n). Some thrombi were detected inside the vascular system (Supplemental Fig.5o).

Furthermore, Endothelin 1/2/3, a protein involved in blood-vessel vasoconstriction, was highly expressed in the first phase patients but reduced in others (Supplemental Fig.5q-t). The triad, angiotensin II, activated mast cells, and activated macrophages can increase vascular permeability (31,53,54).

Concerning the study limitation, we should highlight important points of the current study. Notably, the chronology of testicular pathogeny was based on the presence of advanced germ cells in testis parenchyma and the time of ICU admission. Although our histological and molecular alterations reinforced this conceptual classification, the data of all individuals were also presented in a single group. Moreover, only severely ill patients, who died from COVID-19, were included in the study. We should mention that recent data on semen demonstrate that patients recovered from COVID-19 reestablish their sperm quality after three months of the infection(54). Not all COVID-19 patients studied presented the same comorbidities, and Controls were not submitted to the medications used for COVID-19 patients. On the other hand, we managed to harvest and process the testicles on the same day as the death occurred, in contrast with many cadaver studies with long organ collection delays, which may have compromised the precise histology, detection of virion particles, and perception of peptides, mRNA and hormones fluctuations.

## CONCLUSIONS

Herein, we used different and sensitive methods to detect SARS-CoV-2 proteins, RNA, and virus particles in the testis. In figure 7, we hypothesized the potential viral, cellular, and molecular mechanisms of infection and damage by SARS-CoV-2 in testes of non-vaccinated and severely ill patients. A direct influence of SARS-CoV-2 in testicular cells might deregulate ACE2, elevating the levels of angiotensin II, a potent pro-inflammatory and angiogenic peptide. Angiogenic and inflammatory factors might induce the infiltration and activation of mast cells and macrophages. Therefore, the deleterious effects evidenced in the testes of COVID-19 patients, i. e. fibrosis, vascular alteration, inflammation, tunica propria thickening, Sertoli cell barrier loss, germ cell apoptosis, and inhibition of Leydig cells, may be associated with elevated angiotensin II and activation responses of mast cells and macrophages (Fig. 7a). A protein-protein (or gene-gene) network constructed from the STRING database (Fig. 7b) shows the interaction of genes associated with angiotensin II regulation, activation of mast cells and macrophages, and testis tubular and intertubular compartment alterations.

Our multidisciplinary findings might contribute to a better understanding of SARS-CoV-2 tropism, biology, and impact on testes and male fertility. This is the first study that shows: 1) the high SARS-CoV-2 tropism to the testis; 2) one mode of SARS-CoV-2 entrance in testes; 3) SARS-CoV-2 preferred infection and replication in spermatogonia and macrophages; 4) that the virus remains infective after a long infection period in the testes (viral reservoir); 5) high levels of angiotensin II and activated mast cells and macrophages are critical players in promoting all testicular alterations; 6) the more extended severe condition, the lower the number of surviving germ cells; 7) fluctuation in several essential testicular genes; 8) that the intratesticular testosterone levels are 30 times reduced in testes of COVID-19 patients; 9) the prevalent types of collagen present in SARS-CoV-2 mediated testicular fibrosis; 10) the fluctuation of vasoconstrictive peptides in testes of COVID-19 critically ill patients.

## Data Availability

All data produced in the present study are available upon reasonable request to the authors

## ACKNOWLEDGEMENTS

This work was primarily financed by Ferring COVID-19 Investigational Grant (grant n° FIN0042393, to G.M.J.C). G.M.J.C. also received resources from FAPEMIG (APQ-01078-21). Other specific grants from coauthors punctually helped in some experiments and logistics of the present study. AFV was supported by a FAPESP (grant #18/17647-0) when the study was conducted. GRFC is supported by a FAPESP (grant #20/07419-0). M.H.F. was supported by the Laboratório São Paulo/BH-MG. M.L.N. is supported by FAPESP (grant # 2019/07250-9 and #2020/04836-0) and CNPq. F.G.F. received grants from FAPEMIG (CBB-APQ-03081-17, and CBB-APQ-04295-17), and from Rede Mineira de Pesquisa e Inovação para Bioengenharia de Nanossistemas (RM PI-BEM – FAPEMIG - TEC - RED-00282-16). R.S.A was supported by CNPq (R.S.A.: 312688/2017-2 and 439119/2018-9). We thank Dr. Luiz Orlando Ladeira, Dr. Rodrigo Ribeiro Resende, Dr. Ary Correa Jr, and Dr. Iara Borges for their help and gently allowing the use of their Lab equipment. We also thank the Image Acquisition and Processing Center (CAPI-ICB/UFMG) and the Centro de Microscopia da UFMG (CM-UFMG) for their technical assistance. We especially thank the families of the deceased COVID-19 patients for understanding the importance of our study.

## AUTHOR CONTRIBUTIONS

Study design: G.M.J.C., G.D.C., A.K.Z., L.G.G.P., M.H.F. Performed experiments: G.M.J.C., S.M.S.N.L, A.F.A.F., N.T.W., M.R.B., G.H.C.S., A.F.V., L.M.A., F.R.S., A.L.C.B., G.R.F.C; E.M.N.M, R.S.A., L.M.K., G.R.F.C, E.M.N.M., L.A.M., C.B., P.R, Y.L.G. Data analysis: G.M.J.C., S.M.S.N.L, M.R.B., A.F.A.F., N.T.W., G.H.C.S., A.F.V, L.M.A., G.D.C., H.C.G., V.V.C., F.G.F., M.L.N., M.B.E., A.M.M., K.W.O., B.P.M., M.S.P., T.P.F., M.H.F. Intellectual, technical and resource support: G.M.J.C., G.D.C., H.C.G., V.V.C., F.G.F., M.L.N., R.S.A, G.B.M., A.K.Z, M.H.F. Wrote the first draft of the paper: G.M.J.C., A.K.Z., M.H.F. All authors reviewed and approved the final version of the manuscript.

## ADDITIONAL INFORMATION

Correspondence and requests for materials should be addressed to Guilherme M. J. Costa.

## SUPPLEMENTAL FIGURES

**Supplemental Fig. 1.**
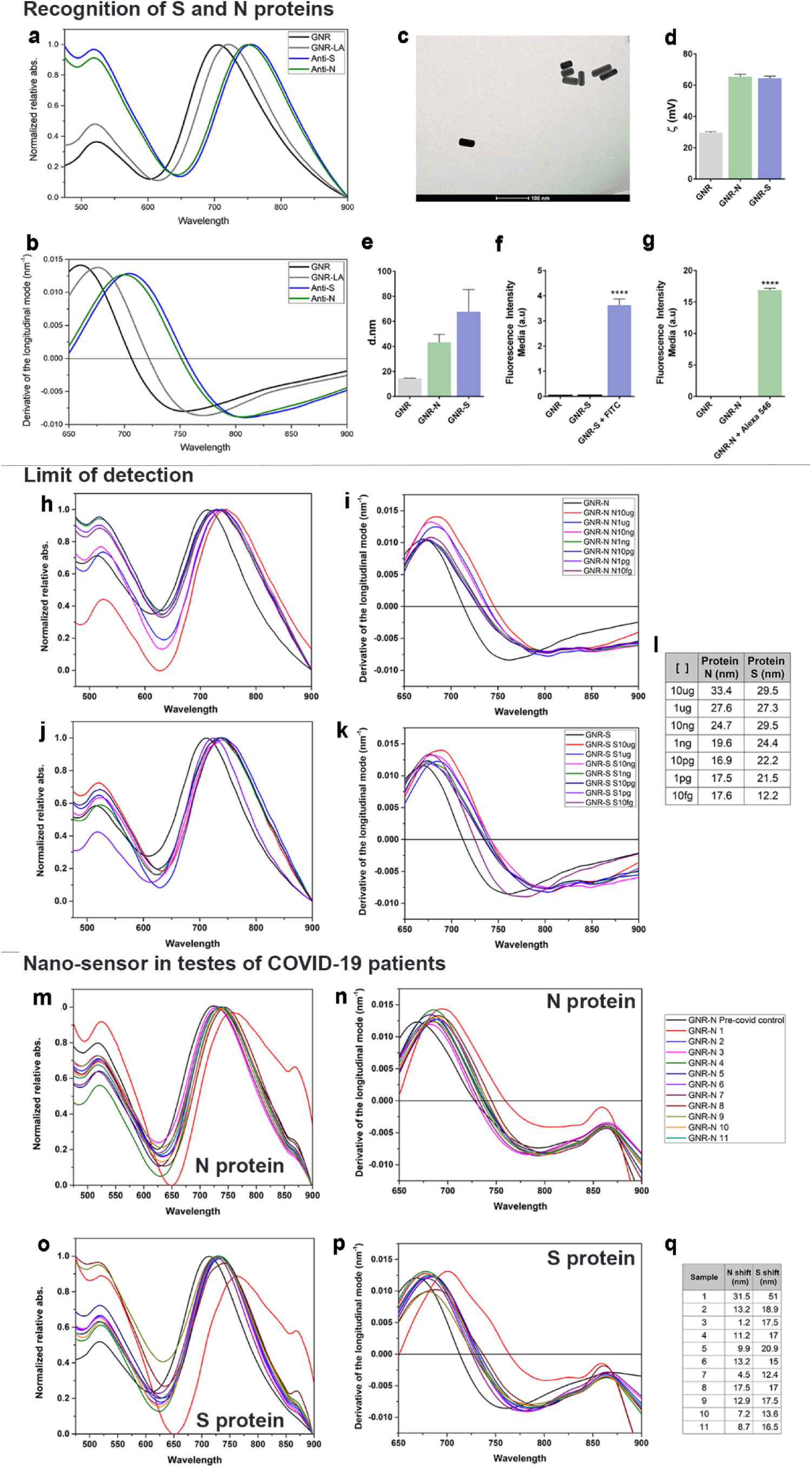
COVID-19 nanosensor platforms. Gold nanorods (GNRs) were coated with anti-S and anti-N antibodies to construct a COVID-19 nanosensor and then characterized by different techniques. a) UV-Vis LSPR spectra of bare GNRs, lipoic-acid coated surface (GNR-LA) and GNR nanosensors recognising S- and N-proteins, respectively. b) derivative curve of the observed light absorbance red shifts. c) TEM image of the GNRs. d) stability of both nanosensors by zeta potential measurements. e) the dynamic light scattering of both nanosensors and their respective hydrodynamic radius. f-g) fluorophores binding both nanosensors. h-k) Light absorbance spectra due to LSPR and respective derivative curves regarding the limits of detection of both nanosensors exposed to different concentrations of S- and N-proteins in means of nm shifting. m-p) Light absorbance spectra due to LSPR and respective derivative curves of the nanosensors recognising the presence of SARS-CoV-2 in each sample compared to non-infected Control. q) the LSPR shift (nm) of each nanosensor after samples exposure. (mean ± SD, ****p<0.001)

**Supplemental Fig. 2.**
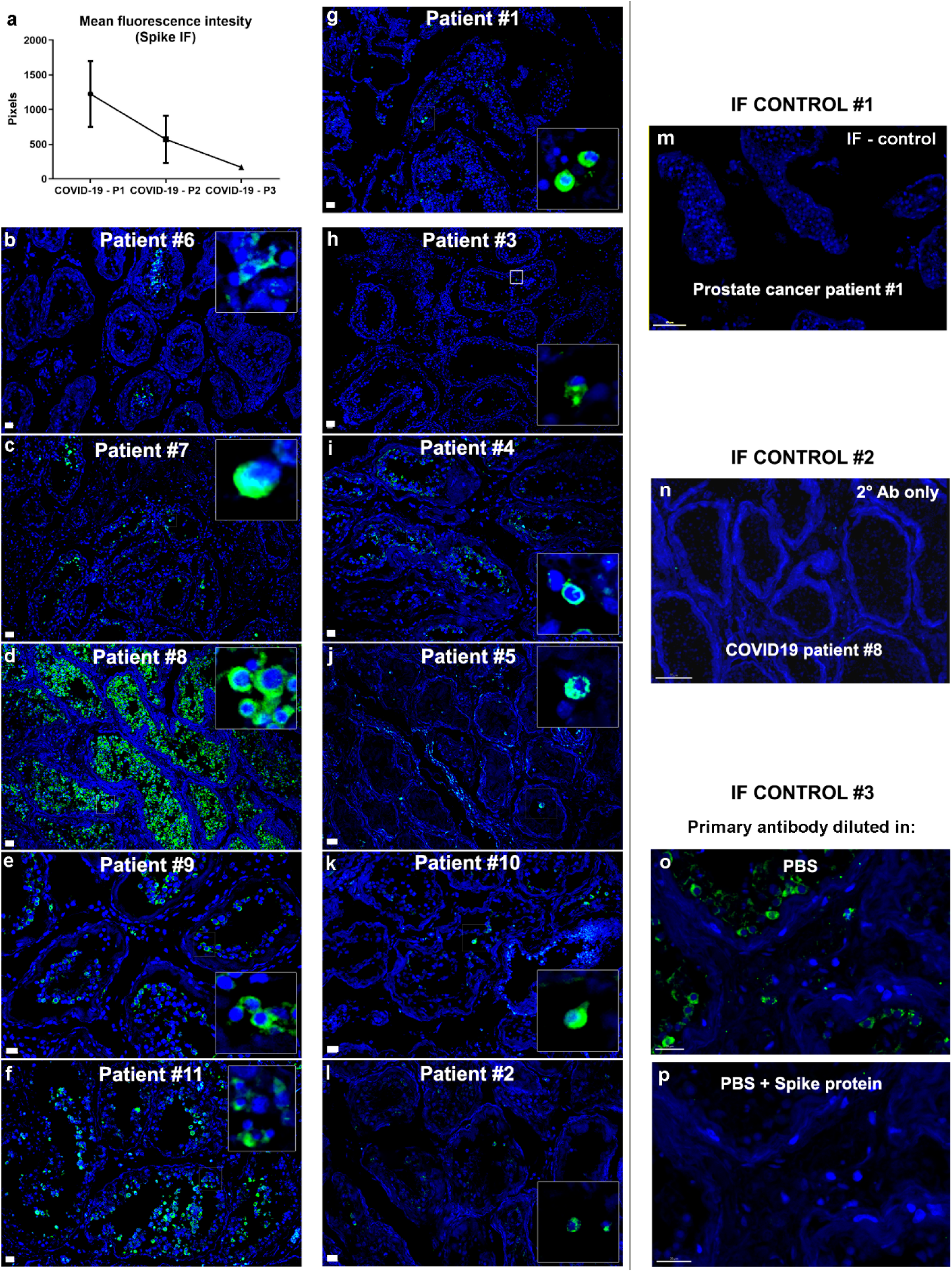
Immunofluorescence against S-protein in testes of all COVID- 19 patients. a) mean fluorescence index (in pixels) evaluating the large images of immunostainings against S-protein. b-l) images of testis parenchyma of COVID-19 patients. b-f) first phase COVID-19 patients. g-k) second phase COVID-19 patients. l) third phase COVID-19 patient. Inserts depict labeled germ cells in all patients. Scale bars = 20 µm. m-o) Controls of the immunofluorescence reactions. m) negative image of Control patient #1 (Scale bar = 70 µm). n) negative control, omitting the primary antibody. o-p) antigen control, comparing the tissue incubated with primary antibody (o) and primary antibody previously incubated with purified Spike protein.

**Supplemental Fig. 3.**
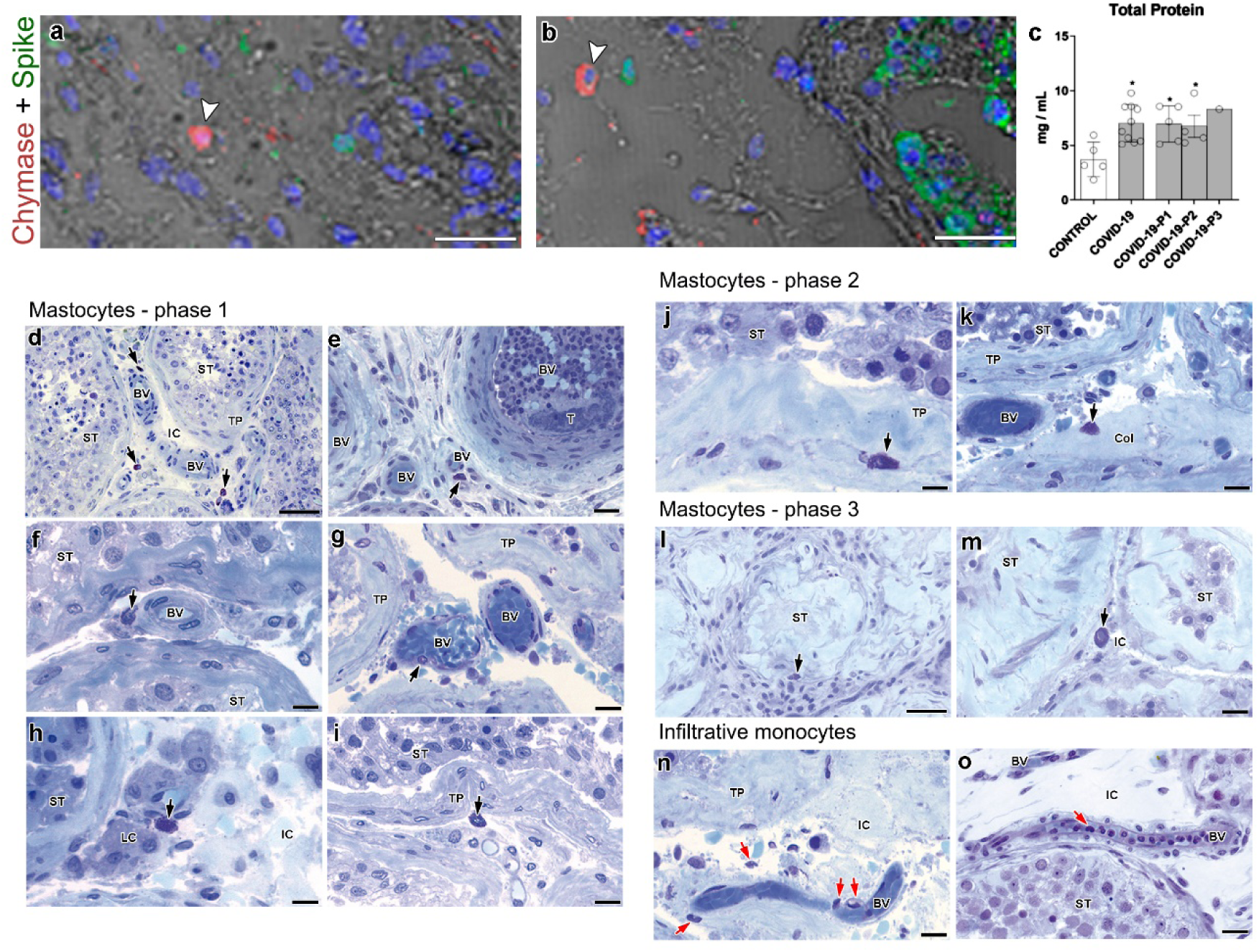
Histology of activated mast cells and infiltrative monocytes in COVID-19 patients. a-b) activated mast cells (chymase +, white arrowheads) are not labeled for the S-protein (green). Blue = dapi staining. (Scale bars = 50 µm). c) quantity of protein per milligram of tissue indicating an inflammatory process (t-test; two tailed; infected groups vs control, *p<0.05). d-m) high number of mast cells in testes of COVID- 19 patients (arrows) (Scale bar = 50 µm). d-e) mast cells near blood vessels (BV) filled with immune cells (T represents a thrombotic area) (Scale bar = 20 µm). f-g) perivascular mast cells (arrows) near intact (f) and disrupted (g) blood vessels (Scale bars = 15 µm). h) mast cell (arrow) near Leydig cells (Scale bar = 15 µm). i) mast cell (arrow) near thickened tunica propria (Scale bar = 15 µm). j) mast cell (arrow) inside the seminiferous tubule (Scale bar = 15 µm). k) mast cell (arrow) near a collagen matrix (Col) in the intertubular space (Scale bar = 15 µm). l) mast cell (arrow) inside a fibrotic seminiferous tubule (Scale bar = 50 µm). m) mast cell in the intertubular area of a third phase patient (Scale bar = 25 µm). n-o) high number of infiltrative monocytes (red arrows) in the intertubular compartment (IC) (Scale bars = 20 µm).

**Supplemental Fig. 4.**
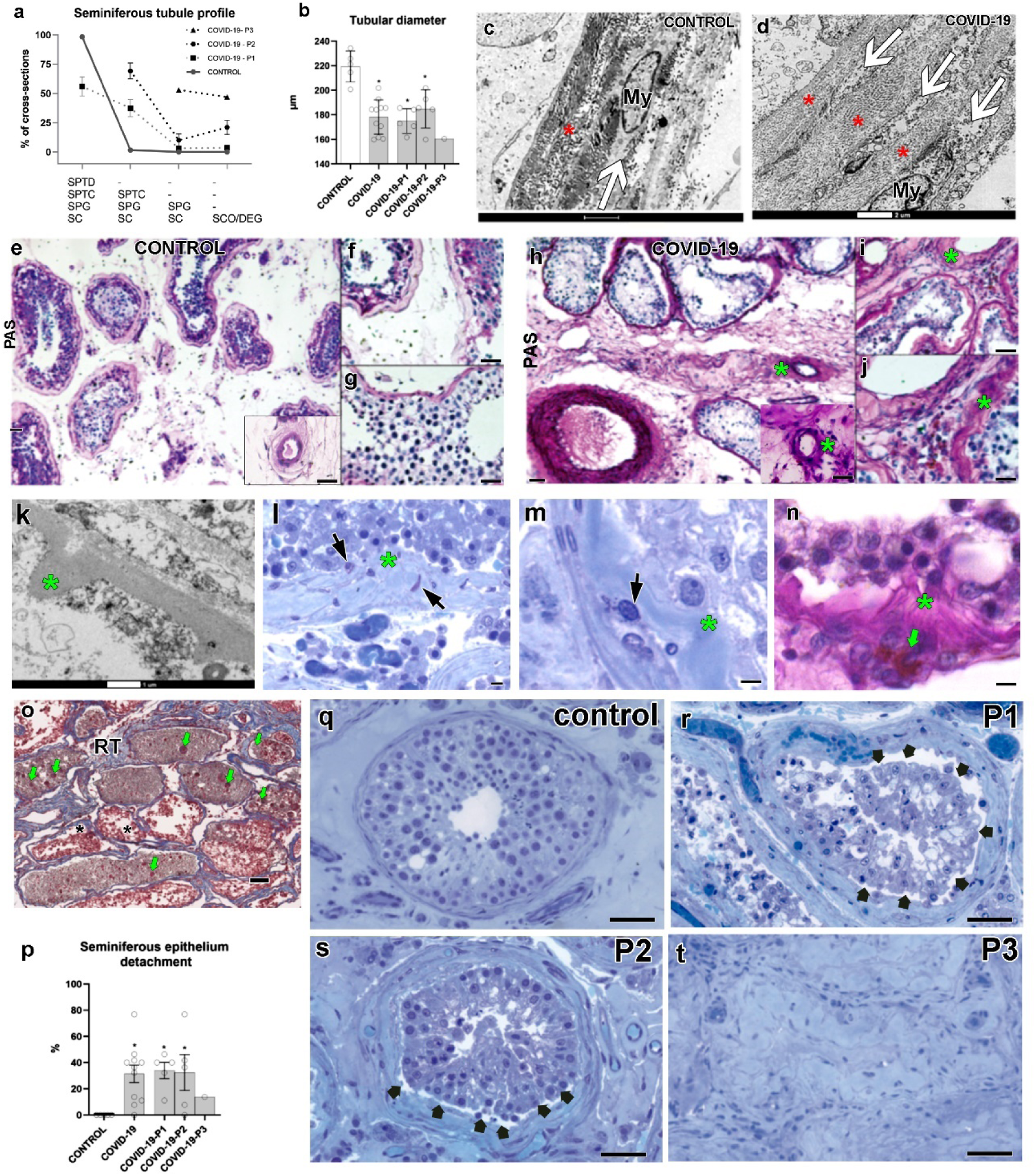
Tubular compartment morphological alterations. a) quantification of the seminiferous tubule cross-sections profile (in %) in Controls and COVID-19 patients. b) seminiferous tubule diameter in Controls and COVID-19 patients (t-test; two tailed; infected groups vs control, *p<0.05). c-d) high number of collagen fibers (asterisks) and peritubular myoid cells (My, white arrows) in COVID-19 patients (Scale bars = 2 µm). e-j) basement membrane (PAS+) in Controls (e-g) and COVID-19 patients (h-j) (Scale bars = c: 30 µm). Asterisks denote high deposition of glycoproteins in the basement membrane and surrounding blood vessels. k-n) convoluted appearance of the basement membrane of COVID-19 patients (asterisks) (Scale bars = s: 1 µm; t-v: 10 µm). Mast cells (black arrows) and macrophages (green arrows) were observed near these areas. o) rete testis (RT) area filled with sloughing and apoptotic germ cells (green arrows) in COVID-19 patients (Scale bar = 50 µm). p-t) seminiferous epithelium detachment (black arrows) from the tunica propria in Controls and COVID-19 patients (t-test; two tailed; infected groups vs control, *p<0.05) (Scale bars = 50 µm).

**Supplemental Fig. 5.**
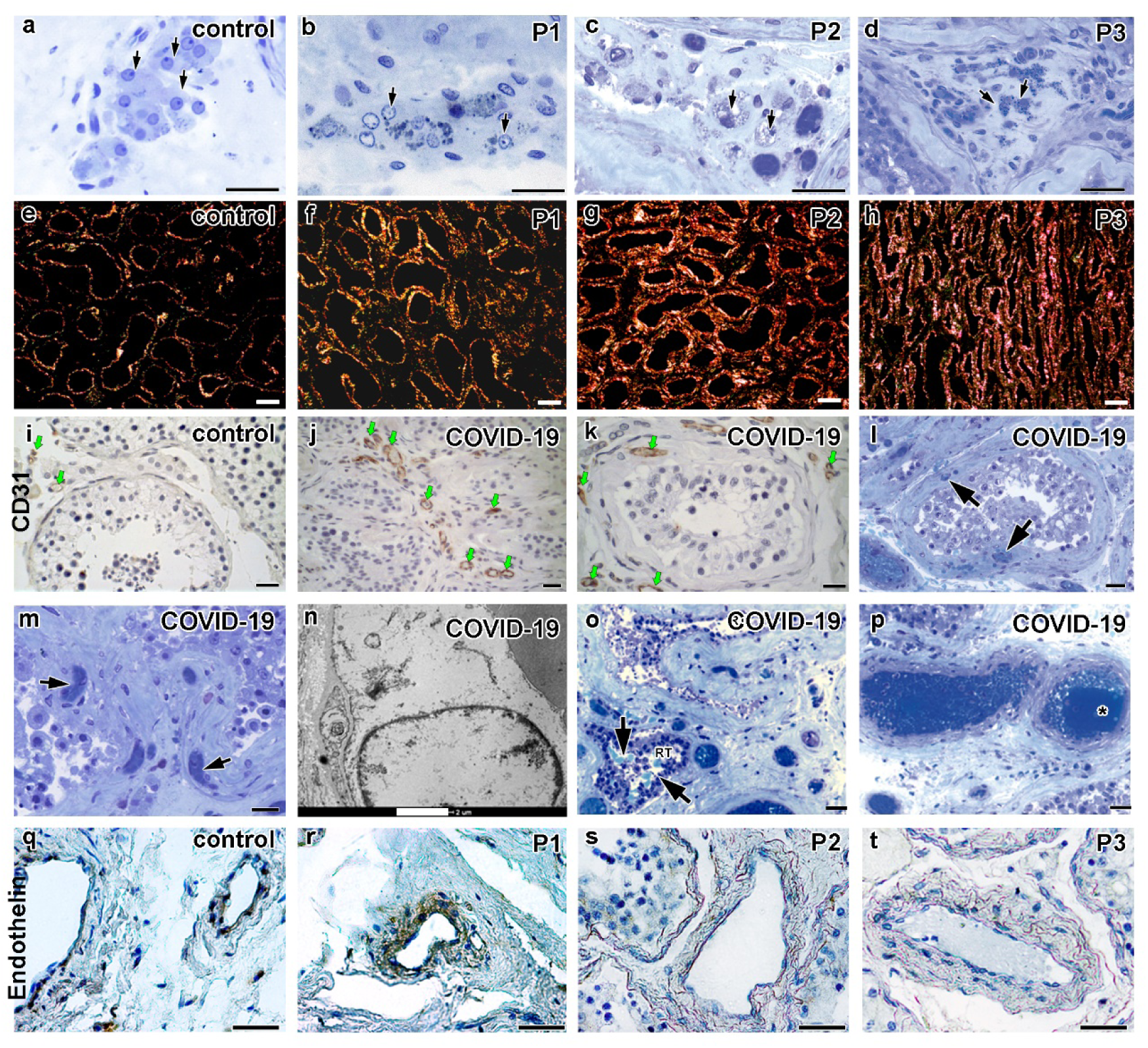
Leydig cell, collagen deposition, and blood vessel alterations in COVID-19 patients. a-d) Leydig cell morphology in Controls and COVID-19 patients (Scale bars = 30 µm). b-d) vacuoles and granules in the Leydig cell cytoplasm. e-h) Picrossirius staining demonstrating Birefringence in Yellow Orange (type I collagen) and green (type III collagen) (Scale bars = 100 µm). i-k) CD31 immunolabeling in Controls (i) and COVID-19 patients (j-k) evidencing the newly formed blood vessels in testis parenchyma (Scale bar = 30 µm). l-m) blood vessels inside tunica propria (arrows) of COVID-19 patients (Scale bars = 30 µm). n) TEM image of an immature endothelial cell identified in tunica propria (Scale bar = 2 µm). o) red blood cells (arrows) inside the rete testis (RT) lumen (Scale bar = 30µm). p) thrombus (*) inside the blood vessel (Scale bar = 30 µm). q-t) Endothelin1/2/3 immunolabeling in Controls (q) and COVID-19 phase one, phase two, and phase three (p1, p2, and p3) patients showing a higher intensity in phase one patients followed by weak labeling in phases two and three patients (Scale bars= 50 µm).

**Supplemental Fig 6.**
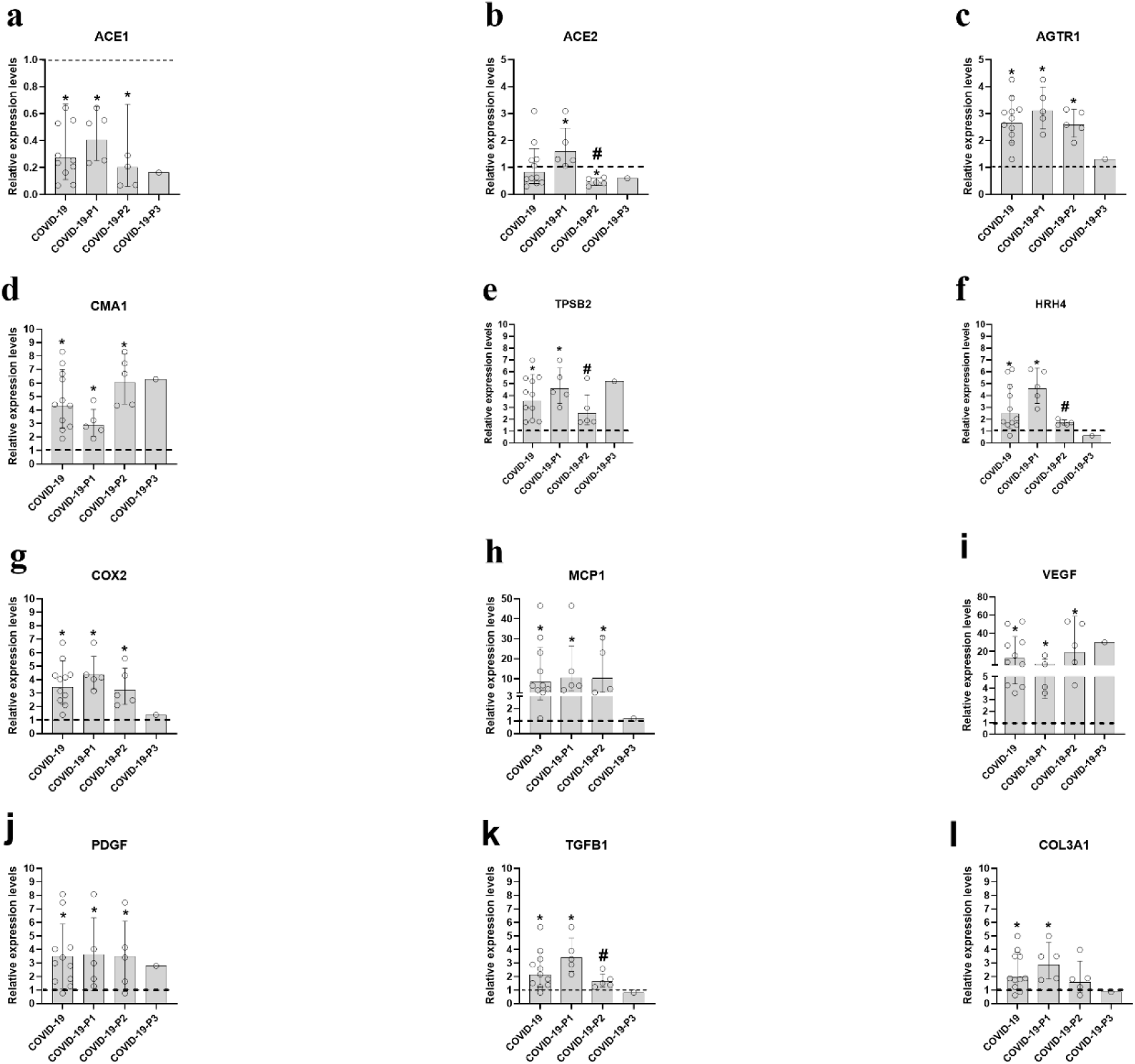
Transcript level of key genes related to high angiotensin II levels, immune cells and vascular system and fibrosis. a-c) Genes from the renin- angiotensin system: ACE1 (a), ACE2 (b), and AGT1R (c). d-e) mast cell genes: chymase (CMA1) (d), and tryptase (TPSB2) (e). f) Histamine receptor (HRH4). g) Cyclooxygenase-2 (COX-2). h) Monocyte chemoattractant protein 1 (MCP1). i) Vascular Endothelial Growth Factor (VEGF). j) Platelet Derived Growth Factor (PDGF). k) Transforming Growth Factor Beta 1 (TGFB1). l) Collagen Type III Alpha 1 Chain (COL3A1). Data are expressed as geometric mean ± SD. *p < 0.05 COVID-19 patients compared to the Control group (which is set at 1; dashed line). # indicates significant differences between COVID-19-P1 and COVID-19-P2 (p < 0.05); t-test (two-sided).

**Supplemental Fig.7.**
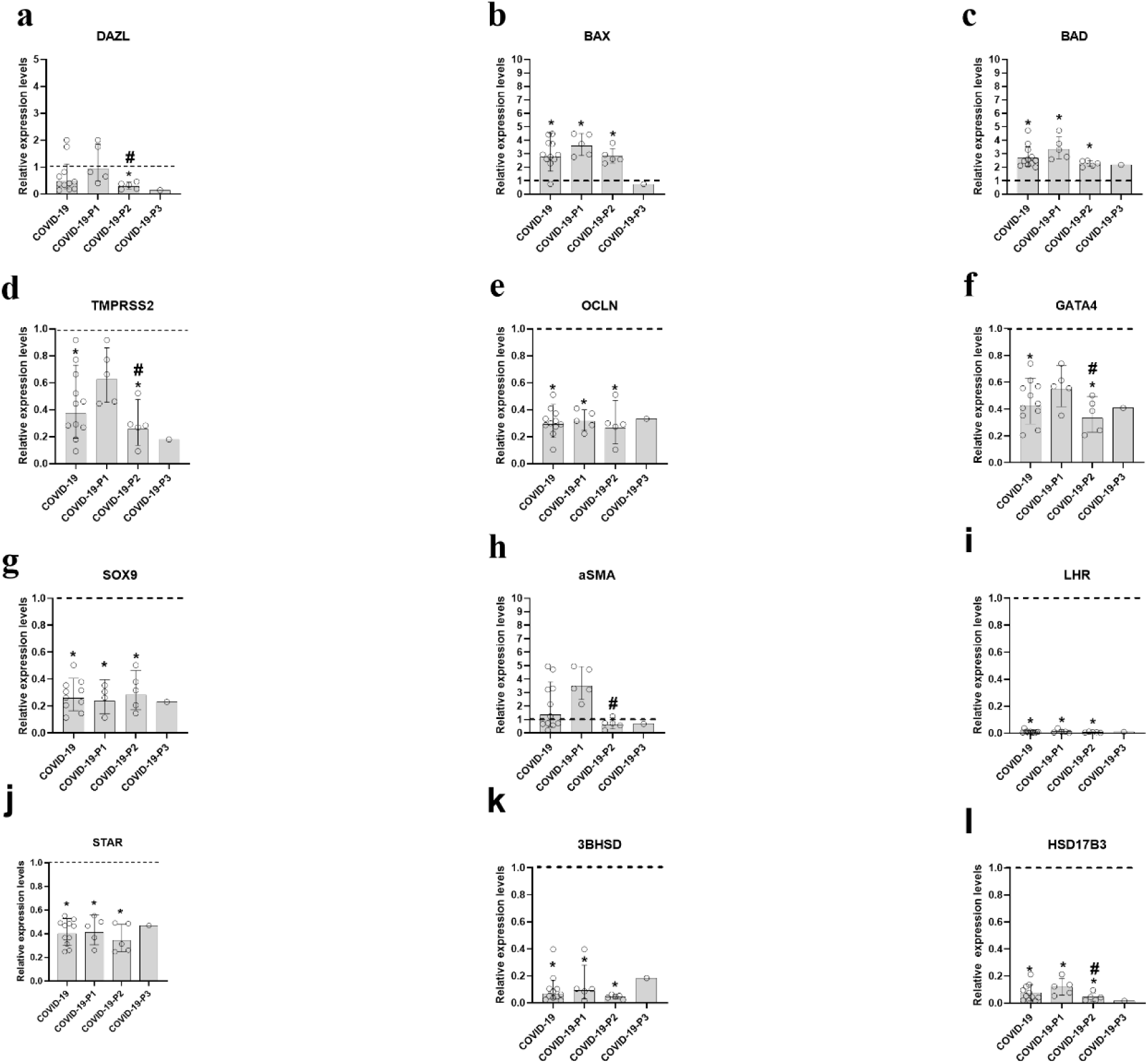
Relative expression of important gene associated to the tubular compartment and Leydig cells. a) Deleted In Azoospermia Like (DAZL). b) BCL2 Associated X Protein (BAX). c) BCL2 Associated Agonist Of Cell Death (BAD). d) Transmembrane Serine Protease 2 (TMPRSS2). e) Occludin (OCLN). f) GATA Binding Protein 4 (GATA4). g) SRY-Box Transcription Factor 9 (SOX9). h) Actin, Alpha Skeletal Muscle (aSMA). i-l) genes related to Leydig cell steroidogenesis: Luteinizing Hormone Receptor (i), Steroidogenic Acute Regulatory Protein (j), 3 Beta-Hydroxysteroid Dehydrogenase (k), Hydroxysteroid 17-Beta Dehydrogenase 3 (l). Data are expressed as geometric mean ± SD. *p < 0.05 COVID-19 patients compared to the Control group (which is set at 1; dashed line). # indicates significant differences between COVID-19-P1 and COVID-19-P2 (p < 0.05); t-test (two-sided).

## TABLES

**Supplemental Table 1.** Clinical data from control patients.

| Control Patient | Age range | Fertility record (children) | FSH (mUI/ML) | LH (mUI/ML) | TTB (ng/dL) | Comorbidity |
| --- | --- | --- | --- | --- | --- | --- |
| #1 | 61-65 | Yes | 2.57 | 3.91 | 227 | Prostate cancer |
| #2 | 56-60 | Yes | 9.44 | 4.42 | 398 | Prostate cancer |
| #3 | 56-60 | Yes | 1.49 | 4,18 | 523 | Prostate cancer |
| #4 | 46-50 | Yes | 1.64 | 3.83 | 646 | Prostate cancer |
| #5 | 56-60 | Yes | 2.41 | 3.68 | 288 | Prostate cancer |
| #6 | 61-65 | Yes | 2.38 | 9.40 | 267 | Prostate cancer |
| **Value of Reference** | - | - | 1.55-9.74 | 0.82-6.22 | 71.8-623 | - |

**Supplemental Table 2.**
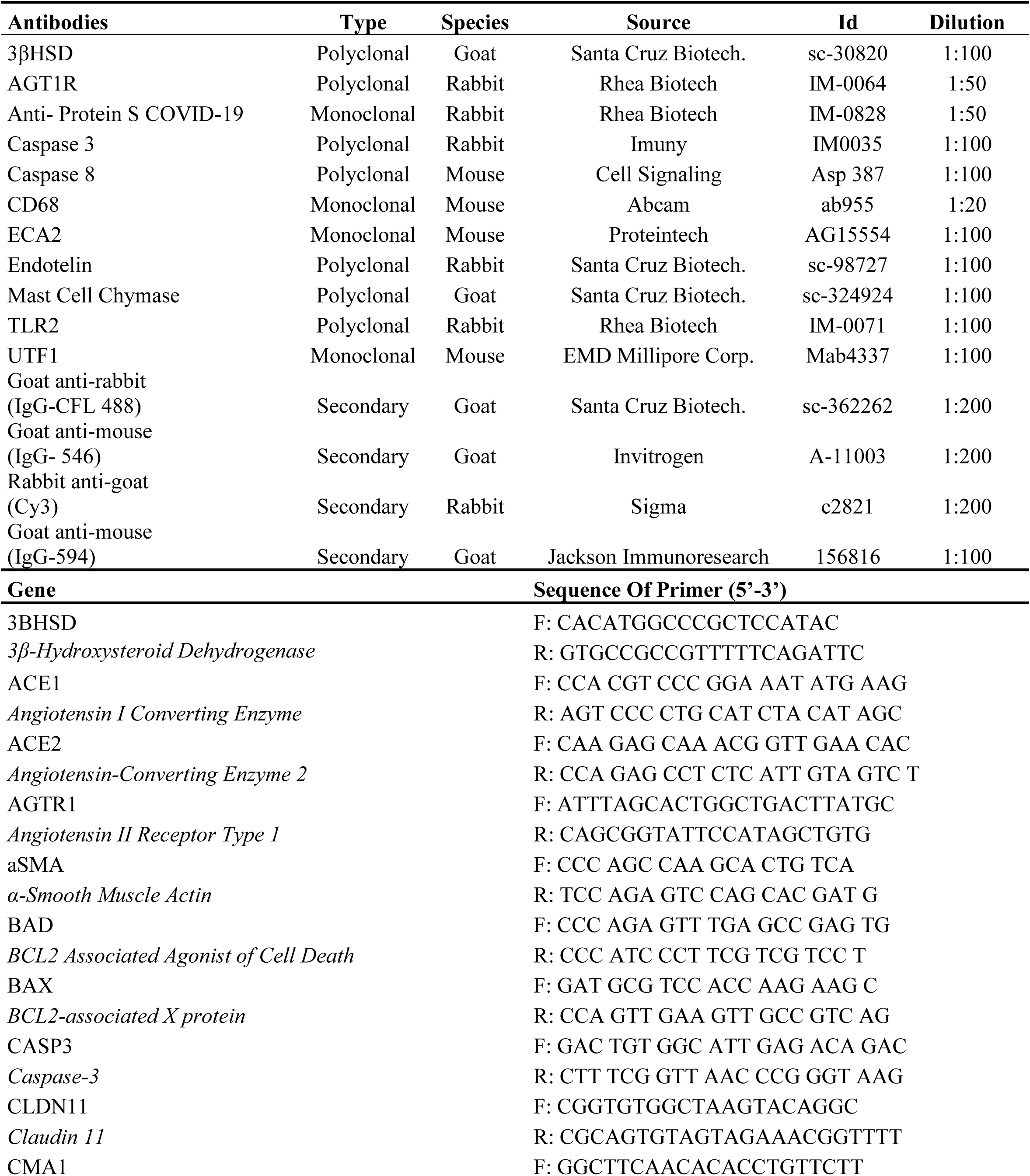
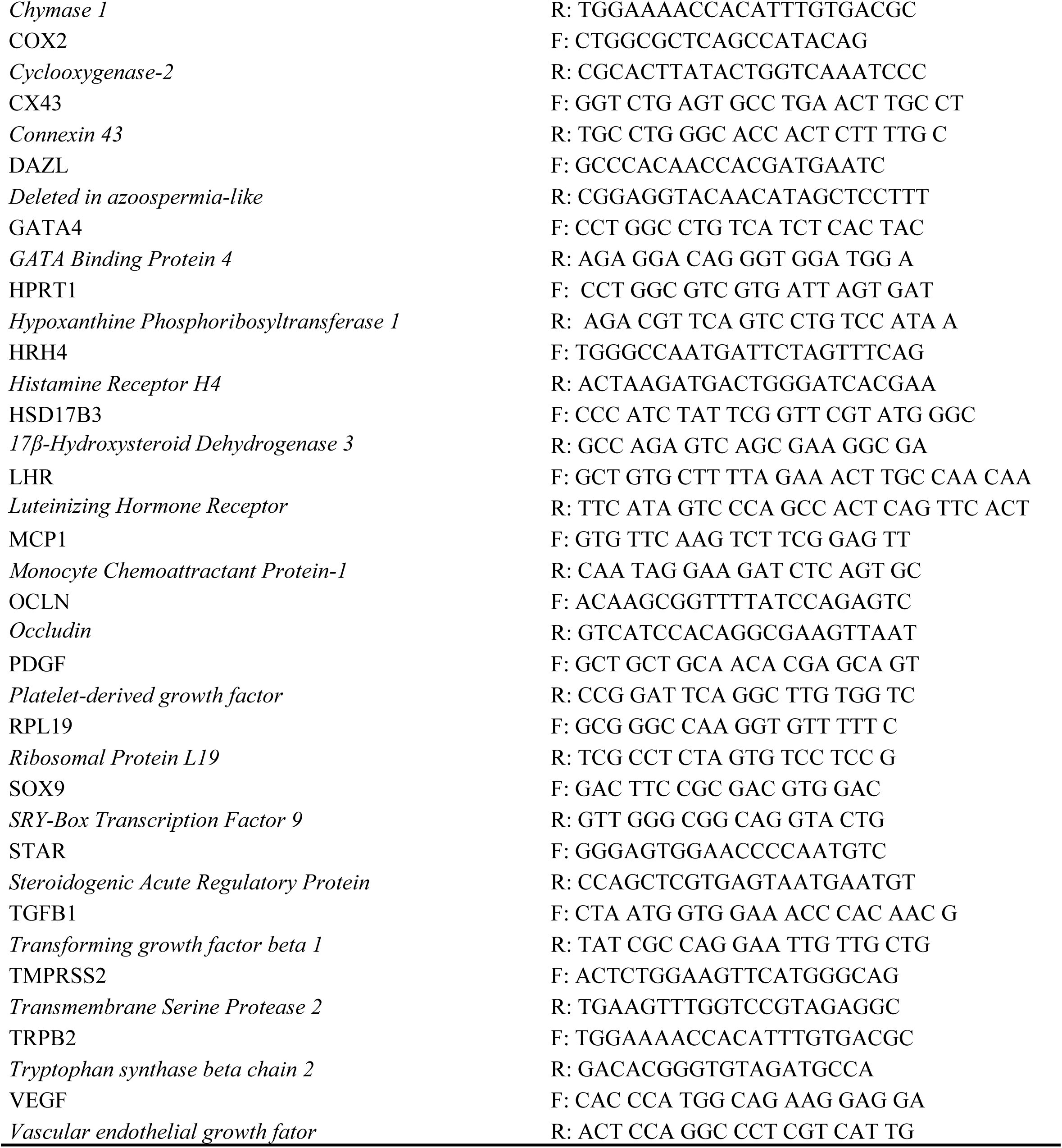
Antibodies and Primers (qPCR) used in this study.

